# The UAE Genome Program: Unique Genetic Insights from 43,608 Individuals

**DOI:** 10.1101/2025.09.12.25334546

**Authors:** Mira Mousa, Roland Michael Olbrich, Inken Wohlers, Amira Al aamri, Aisha Alsuwaidi, Nour al-dain Marzouka, Halima Alnaqbi, Mohamed Salem Alameri, Dymitr Ruta, Jumana Alazazi, Tiago Magalhaes, Joseph Mafofo, Javier Quilez, Mushal Allam, Mohd Saberi Mohamad, Nizar Drou, Youssef Idaghdour, Rifat Hamoudi, Guan Tay, Saleh Ibrahim, Fatima Alkaabi, Asma AlMannaei, Habiba Alsafar

**Author notes:** These authors contributed equally to this work (Mira Mousa, Roland Michael Olbrich).

## Abstract

Here, we present a comprehensive genomic characterization of a cohort of 43,608 Emirati genomes sequenced as part of the Emirati Genome Program (EGP). This study identified more than 421 million single-nucleotide variants and indels and more than 600 million copy-number and structural variants. Small variants had 756 million molecular effects annotated. Of 7.7 million polymorphic variants having an allele frequency (AF) of more than 5% in EGP, 1,348 have a predicted deleterious effect on a protein. Characterization with respect to global variation shows that EGP represents a genetic continuum encompassing the range of African, Asian, and European populations. It is best described by two Arabian, an Eurasian, and an African component, with the predominant Arabian component linked to mitochondrial haplogroups J and T that are commonly attributed to the Middle East. Various aneuploidies of sex chromosomes were detected in 93 individuals overall, and aneuploidy of chromosome 21 was identified in 41 individuals. Median inbreeding coefficient and cumulative runs of homozygosity (ROHs) lengths were increased due to extensive consanguinity, were largest in the groups with Arabian main ancestry components, and were higher than reported for Qatar. Families were identified based on genetic relatedness and classified into 264 families with unrelated parents and 247 families with third- and fourth-degree consanguineous parents. Representative consanguineous pedigrees of families in EGP were outlined. Cumulative ROHs were affected by the main ancestry component and significantly increased in offspring of consanguineous parents, with a pronounced difference between 3rd and 4^th^-degree relatedness. Investigation of cumulative AFs of variants causing Mendelian diseases highlighted genes related to alpha- and beta-thalassemia (*HBB*, *HBA2*) and showed a high burden of variants causing severe recessive diseases, metabolic and retinal disorders, and hearing loss. In summary, EGP represents a landmark effort in characterizing the genetic diversity of the Emirati population, leveraging the largest Middle Eastern cohort reported to date.

## 1 Introduction

The United Arab Emirates (UAE) has actively contributed to global genomic research efforts by addressing the underrepresentation of diverse populations^1,2^. The Emirati Genome Program (EGP) is one of the most extensive single-population cohorts assembled to date, aiming to sequence the entire population of over one million UAE nationals. As of January 2025, 700,000 participants have undergone whole-genome sequencing (WGS) on one of three high-throughput sequencing platforms (Illumina, MGI, and Oxford Nanopore Technologies). This diverse, multiplatform approach enables the comprehensive collection of a wide range of genetic information.

Generating a population-specific reference database is pivotal to establishing an advanced healthcare system that supports targeted research, preventive screening applications, and the integration of precision medicine. The human genomic architecture displays substantial variability across diverse populations due to the intricate interplay of genetic drift, migration, selection pressures, and local adaptation^3,4^. These genetic signatures impact disease susceptibility, pharmacogenomics, and population health^5^. However, most genetic studies, which are primarily focused on European populations, are generally lacking in generalizability, with an underrepresentation of Middle Eastern populations, underscoring the need for more inclusive research.

Collectively, Middle Eastern populations exhibit a high degree of genetic heterogeneity and population diversification, influenced by the convergence of diverse ancestral lineages, including those of Arabian, Persian, Turkish, and Levantine populations^6,7^. Furthermore, high degrees of consanguinity due to cultural practices in some Middle Eastern societies have led to an increased prevalence of runs of homozygosity (ROHs) in these populations, which influences the occurrence of recessive genetic disorders^8–10^. The scarcity of population reference datasets for Middle Eastern populations has constrained the exploration of genetic diversity and the underlying genetic basis of various diseases. This can be mitigated by documenting population-specific reference alleles, which will facilitate the examination of disease associations with variants and sequences that are distinctive or prevalent in Arab populations.

Previous studies in the UAE have investigated population genetic diversity using whole-genome sequencing (WGS) and whole-exome sequencing (WES) datasets of 153 and 1,000 individuals, respectively, aligned to the widely utilized Genome Reference Consortium (GRC; GRCh37) reference genome. The EGP variome of 43,608 individuals reported here is the largest cohort of a single Middle Eastern population examined to date, comprehensively characterizing the genetic makeup of the Emirati population in terms of small and structural variations, chromosomal aberrations, UAE common and specific variations, familial relationships, and homozygosity. Furthermore, we describe ancestry components that characterize the Emirati population and evaluate their influence on genetic factors.

## 2 Materials and Methods

### 2.1 Sample collection

As part of the EGP Consortium, participant enrollment, sequencing, data storage, and preprocessing were implemented by M42 Healthcare (https://m42.ae). Recruitment was conducted across various sites, including operational centers, Abu Dhabi National Exhibition Centre halls, community gathering centers known as "Majlis", hospitals, and educational institutions, to ensure comprehensive, representative, and diverse data collection. All participants completed a thorough informed consent process to ensure ethical compliance. The study procedures and protocols adhered to the ethical guidelines and regulations established by the Research Ethics Scientific Committee in the UAE (DOH: DOH/CVDC/2022/1701), Khalifa University (KU: H21-045), and the principles outlined in the Declaration of Helsinki. As part of the stringent security measures surrounding this nationwide effort, access to personal information and phenotype data is strictly limited. Accordingly, approved research projects will be provided with anonymized data and only with the minimum information necessary for the respective research question. The available participant information in this study is thus limited to age and sex.

For consenting participants, 5 ml of blood was collected. The collected samples were delivered to the M42 OMICs laboratory and associated with the M42 field barcode-generated order. Then, an EGP laboratory barcode was issued and linked for further tracking and identification. Further information on the sample collection is detailed in Supplementary Text 1.1.

### 2.2 DNA extraction and sequencing

The blood fractionation process involved using a Hamilton easyBlood Star for autofractionation. The DNA extraction process was performed with the Chemagic 360D Automated DNA Extraction Kit (Perkin Elmer) and the Chemagic DNA Buffy Coat 200 Kit H96. Quality control measures were implemented using a Nanodrop 1000 spectrophotometer (Thermo Fisher) for the determination of A260/230 and A260/280 ratios, with DNA concentration quantified using the Qubit BR Assay Kit from Invitrogen on a Fluoroskan Plate Reader (Thermo Fisher). The extracted DNA was divided into two aliquots and placed into matrix tubes (Thermo Fisher), one for immediate working storage and the other for biobanking.

After DNA quality control, genomic DNA normalization was performed on an OT-2 Liquid handler (Opentrons), followed by library preparation on the Tecan Freedom Evo 200 NGS Platform using the Illumina DNA PCR-free Library Prep Kit. Library barcoding was achieved by incorporating IDT® for Illumina® UMI DNA/RNA UD Indexes Set A, with subsequent library quality control using the Qubit ssDNA Assay Kit on the Qubit 4.0 (ThermoFisher). Sequencing was performed using the NovaSeq 6000 S4 Reagent Kit v1.5 (300 cycles) on the NovaSeq 6000 System, with BCL files generated and demultiplexed using bcl2fastq on the Dynamic Read Analysis for GENomics (DRAGEN) platform by Illumina. All the samples were sequenced to obtain 90 gigabases (Gb) of raw data. For good-quality whole-genome sequences and a high mapping rate, 90 Gb translates into approximately 30X effective coverage, the target coverage of the EGP. Samples below this threshold were subjected to a top-up procedure, as described in Supplementary Text 1.2.

### 2.3 Data processing, variant calling, and variant quality control using DRAGEN

The DRAGEN Bio-IT Platform version 3.9 was used to perform the initial steps in the bioinformatics analysis pipeline. These steps included demultiplexing and base-calling of BCL files, as well as the generation of single-sample FASTQ files, including adapter trimming. The resulting FASTQ files were passed to the DRAGEN Germline Pipeline. This process involved aligning sequence reads to the GRCh38 reference genome and incorporating several critical analytical components, including sorting, duplicate marking, base quality score recalibration, variant calling, and the computation of comprehensive summary statistics.

In addition to primary variant calling, the DRAGEN pipeline has yielded diverse supplementary outputs, offering valuable insights into various aspects of genomic variation and structural complexity. Specifically, the pipeline generated files in standard VCF file formats for structural variants (SVs) using Manta^11^ as a caller, and for small variants and copy number variants (CNVs) using its own callers. Furthermore, DRAGEN estimates ploidy and generates a BED file for regions of homology (ROHs). Furthermore, the analytical results were accompanied by metric files that succinctly summarized each sample’s findings and statistics, enhancing the comprehensiveness and interpretability of the generated data. The DRAGEN pipeline has been intensively benchmarked^12^ and is comprehensively documented at https://support-docs.illumina.com/SW/DRAGEN_v39/Content/SW/DRAGEN/Software.htm.

### 2.4 Annotation of variants

BCFtools^13^ facilitated the decomposition of multiallelic variants into distinct rows, alongside executing operations for left alignment and normalization. Subsequently, the command line version of Variant Effect Predictor (VEP) (v110.1)^14^ was employed with the ‘--everything’ flag for comprehensive variant annotation using the following GRCh38-specific databases: Ensembl (release 110.584a8f3)^15^, 1000 Genomes (phase3)^16^, COSMIC (release 97)^17^, ClinVar^18^ (as of January 2023), HGMD-PUBLIC (v20204)^19^, dbSNP (release 154)^20,21^, Genecode (version 44)^22^, gnomadADg (v4.1)^23^ 38, PolyPhen (version 2.2.3)^24^, RegBuild (version 1.0), and SIFT (version 6.2.1)^25^. The filter_vep^14^ function was used to identify genetic variants (SNP or indel) that have not been previously documented in the annotated dbSNP, gnomAD Exomes, and Genomes, or the 1000 Genomes versions. Repeat-region variants were defined as those located within regions designated by the UCSC RepeatMasker track^26^. This step was taken to identify variants in repetitive genomic regions, which are prone to alignment artifacts and low-confidence variant calls. For the identification of variants that are specific to the EGP cohort, allele frequencies (AFs) were also checked in the BRAVO database of 705 million variants observed in 132,345 genomes sequenced as part of the TOPMed^27^ project.

For the analysis of predicted loss-of-function (pLoF) variants from the variome dataset, we considered variants with high impact (consequences: splice acceptor variant, splice donor variant, stop gained, stop lost, frameshift variant, start lost, transcript ablation) on a canonical Ensembl transcript, as per VEP. Variants overlapping repetitive regions were excluded based on RepeatMasker annotations. Following the example of Salheen et al.^28^, the remaining variants were filtered for a minor allele frequency smaller than 1%, eliminating common and polymorphic ones, and further restricted the dataset to variants classified as pathogenic (P) or likely pathogenic (LP) (P/LP in the following) according to ACMG annotation obtained using the GeneBe tool (GeneBeClient-0.1.0-a.9).

### 2.5 Mitochondrial haplogroups, ploidy, and ROH annotation

Mitochondrial variants were called by DRAGEN^12^, and mitochondrial haplogroups were determined using the stand-alone version of Haplogrep3^29^. The results generated by DRAGEN’s ploidy caller offer information on the ploidy and sex karyotype of each sample, excluding information on the mitochondrial DNA, unlocalized or unplaced sequences, alternate contigs, decoy contigs, and Epstein–Barr virus sequences. DRAGEN quantifies ploidy as the median coverage of the respective chromosome divided by the median autosomal coverage.

The most frequent and infrequent ROHs were determined based on the count of samples sharing the same region (considering a 10 kb window size) as calculated by the kpPlotDensity function from the karyoploteR^30^ package in R. The 95% (8,029 samples) and 5% (1,500 samples) quantiles were used to define the cutoffs for the highest- and lowest-frequency windows, respectively, ensuring a balanced length of the frequent and infrequent ROH regions. Adjacent windows were merged using the reduce function from the GenomicRanges^31^ package in R. Only autosomes were included in the ROH analysis, with centromere regions excluded. The plotKaryotype function was used to visualize the results. To assess the overlap of the frequent and infrequent ROHs with autosomal dominant (AD) and autosomal recessive (AR) genes, we used a list of genes with definitive gene-disease relationships, as classified by inheritance patterns in the ClinGen^32^ database (accessed December 5, 2024).

The ROHs were classified into three categories according to length: short (< 14.73 Mb), medium (>=14.73 Mb and < 63.03 Mb), and long (>= 63.03 Mb). The thresholds for these categories were determined by unsupervised clustering using a three-group Gaussian model through the mclust^33^ package in R.

### 2.6 Population structure analyses

For admixture analyses, we used a set of previously compiled and quality-controlled global population-informative variants from 12 studies, encompassing 127,261 variants observed in 5,429 individuals from 144 diverse populations^34^, which are based on various sources, e.g., ^6,7,16,35^. Using the supervised mode of ADMIXTURE^36^, we computed for each Emirati genome the proportions of continental ancestry for the five global populations of 3,502 individuals as part of the 1000 Genomes Project 3, namely, European (EUR), South Asian (SAS), African (AFR), South American (AMR), and East Asian (EAS) ancestry. The cohort was then stratified into five groups related to these global ancestries by assigning each sample to the group of the highest ancestry component, enabling the identification of underlying genetic substructure within the Emirati population. Furthermore, principal component analysis (PCA) was conducted using smartpca implemented in the Eigensoft^37^ package (v8.0.0) and the population informative variants. PCA was computed for the 1000 Genomes samples, and the samples of the Emirati cohort were projected onto this PCA and vice versa.

### 2.6 Relatedness analyses

The study utilized King software (v2.3.0) to assess familial relationships within the cohort, inferring relatedness up to the 4th degree. This tool integrates algorithms for kinship estimation and identity-by-descent (IBD) segment analysis, providing insights into familial dynamics. Subsequently, a graph was constructed within the R framework based on assigned pairwise relationships to identify familial trios and consanguineous relationships. The graph analysis involved initially creating a subgraph focusing on parent-offspring relationships to identify familial clusters. Clusters were scrutinized to detect families with consanguineously related parents, with relationships inferred up to the 3rd degree between parents. The sex inferred from the DRAGEN ploidy information was utilized to confirm the correct assignment of male and female parents.

### 2.7 Identification and characterization of Mendelian disease variants and genes

We utilized a curated list of Mendelian inherited genes (n = 2,648) and their associated diseases, as reported by Aamer et al.^38^, and refined it using the ClinGen^32^ database to ensure accuracy and consistency. Specifically, we filtered the list to include only genes with a ‘Definitive’ gene-disease category in ClinGen and then examined the AF of P/LP variants from ClinVar^18^ in the UAE cohort. The disease names were standardized according to the ClinGen database, and the variant positions overlapped with frequent and infrequent ROH regions. The cumulative AF of P/LP variants was then summarized at the gene level, disease level, and disease group levels, as categorized by Aamer et al.^38^ Since establishing direct variant-disease links would need manual curation because disease names are not sufficiently standardized on the variant level, we summarize P/LP variants by gene and then use the database’s curated link from genes to standardized disease names.

## 3. Results

### 3.1 The Emirati Genome Program’s large-scale sequencing efforts

A subset of 50,000 participants was selected from the Emirati Genome Project using a uniform random sampling approach without replacement, ensuring that each individual had an equal probability of inclusion and minimizing selection bias. In this subset, 43,608 samples were sequenced in overall 2,232 sequencing runs performed from August 2021 until December 2023 (Supplementary Table 1) and analyzed using the Illumina DRAGEN pipeline, and 6,392 samples were processed in the MGI Ztron pipeline. Investigating the technical variation between the two platforms revealed batch effects related to mean coverage, mapping rate, Q30, total SNPs, and sequencing yield (Supplementary Figure 1) and in the genotypic principal components (Supplementary Figure 2) that led to the exclusion of the MGI dataset in this work. The resulting final dataset comprised 43,608 Illumina sequenced samples that had a median coverage of 36x (range 28x-100x), mapping rate of 98.96%, Q30 ratio of 89.66%, sequencing yield of 128.04 Gb, and a median of 4,115,560 SNVs called (Supplementary Table 1, Supplementary Figure 1). Average alignment coverage and coverage uniformity (Supplementary Figure 3) were investigated for quality control, as well as coverage distribution (Supplementary Figure 4). The estimated fraction of reads in a sample that may be from another source had a median of 0.1% (mean: 0.126%), with all but 27 samples having less than 3% (Supplementary Table 1). The ratio of heterozygous to homozygous variants had a median of 1.76 (IQR: 0.23; ranging between 0.91 and 5.73) (Supplementary Table 1), which closely aligns with previously reported ratios for 1000 Genomes American (1.7) and European (1.6) populations, and is between the ratio observed for East Asian (1.4) and African (2.0) 1000 Genomes (1KG) populations.^39^

Of the cohort’s individuals, 19,015 (44%) are female, and their ages range from 1 to 103 years. The age distribution, stratified by sex, is depicted in Supplementary Figure 5, with the median age of female participants 29 years and of male participants 32 years.

### 3.2 A comprehensive catalog of genetic variation: EGP characterizes the UAE variome

Among the 43,608 individuals in the EGP cohort, 421,605,069 small variants were called, of which 356,964,849 are SNVs (84.7%) and 64,640,220 are indels (15.3%) (Table 1, Figure 1 A), Supplementary Table 2). Of those, 10.0 million had a minor AF larger than 5% and can be considered polymorphic, and 33.4 million had a minor AF of more than or equal to 0.01%, but less than 5%. They can still be considered common, as the allele needs to be observed in a minimum of five individuals to reach this cutoff (Table 1). Most identified variants were either rare (having a minor AF of less than 0.01%, but an allele count larger 1; 248 million small variants (58.83%)) or singletons, representing alleles that are observed only once (130 million (30.86%)), see Table 1. Of SNVs, 2.16% are polymorphic and 7.26% are common (Figure 1 B and Supplementary Table 3). These numbers are higher for indels (3.59% polymorphism, 11.62% common; Figure 1 C); Supplementary Table 4). This is reversed for rare SNVs and indels, here 60.28% of SNVs are rare, but only 50.38% of indels (Figure 1 B) and C); Supplementary Tables 3 and 4). Across the cohort, on average, 5,152,390 small variants were called, with a minimum number of variants per Emirati genome of 4,570,516 and a maximum of 6,799,481 (median 5,113,690; Supplementary Table 1).

**Figure 1.**
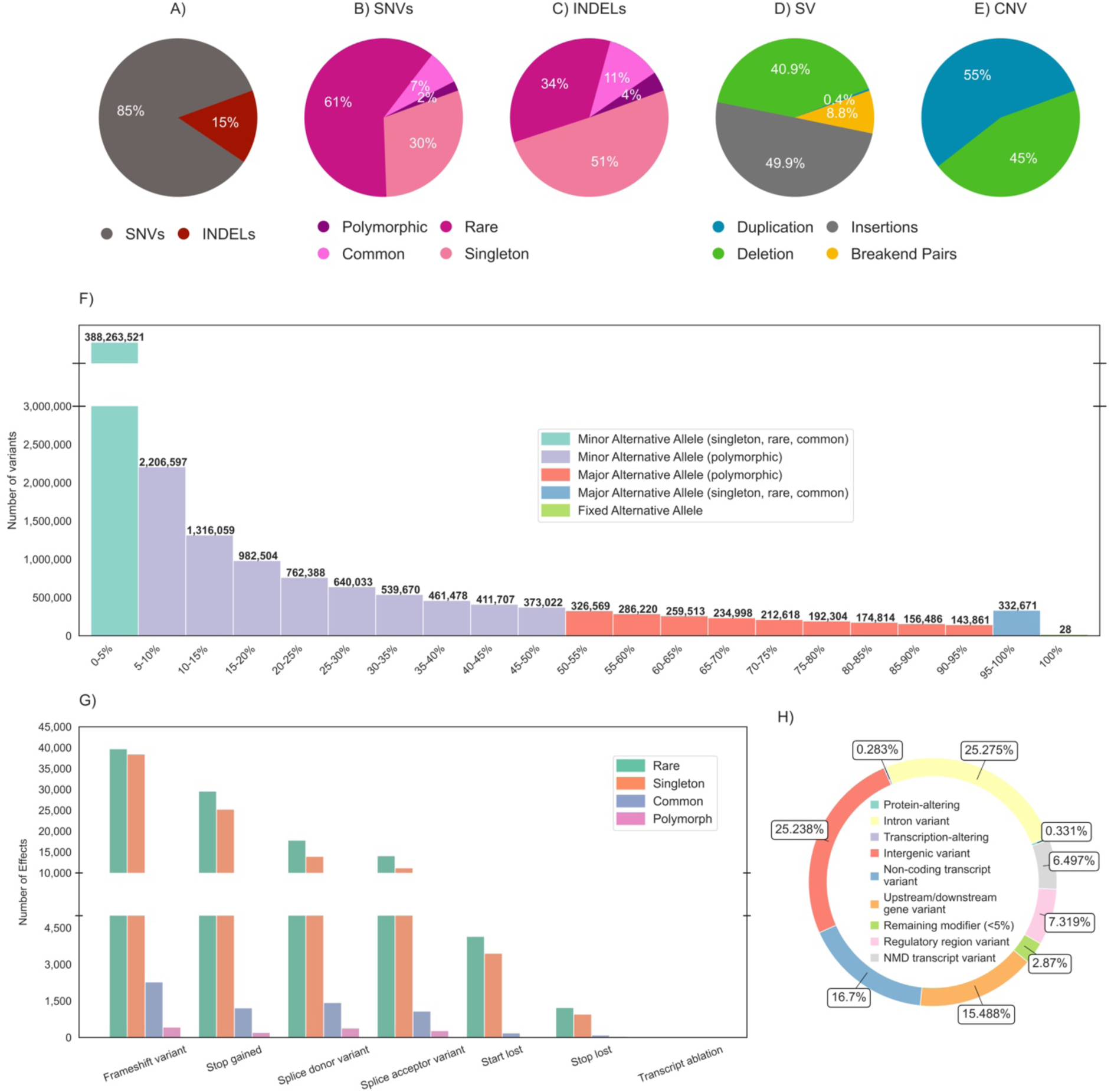
Overview of variant types called in the EGP cohort. A) Distribution of variant types across 421,605,069 small variants (SNVs and indels); prevalence according to allele frequency for SNVs B) and indels C); distribution of variant categories (deletions, insertions, duplications, and break-end pairs) across CNVs D) and SVs E). The histogram F) displays the spectrum of allele frequencies found in SNVs and indel variants identified across 43,608 samples in five percent frequency intervals. Notably, the leftmost bin encapsulates singleton and rare variants, each exhibiting a frequency of five percent or less. In contrast, the rightmost bin shows a set of 28 variants that are present ubiquitously in the population. G) Distribution of variant classes stratified by impact category as per VEP annotation. H) Distribution of selected impact categories in the cohort.

**Table 1.** Variant profile in the cohort. This table presents an overview of the cohort’s 421,649,305 short genetic variants. Common variants represent over 0.1% of all detected alleles in the overall cohort, while rare variants are observed in at least two individuals but less than 0.1%. Singleton variants are unique to a single individual. The breakdown of single-nucleotide variants (SNVs) and short insertions and deletions (indels) across chromosomes is detailed in Supplementary Table 3 and Supplementary Table 4, respectively.

| Variant<br>s | All | Polymorphic |  | Common |  | Rare |  | Singleton |  |
| --- | --- | --- | --- | --- | --- | --- | --- | --- | --- |
|  | [#] | [#] | [%] | [#] | [%] | [#] | [%] | [#] | [%] |
| **All** | 421,605,069 | 10,017,578 | 2.38 | 33,428,152 | 7.93 | 248,033,398 | 58.83 | 130,125,941 | 30.86 |
| **SNVs** | 356,964,849 | 7,698,054 | 2.16 | 25,915,535 | 7.26 | 215,530,807 | 60.38 | 107,820,453 | 30.21 |
| **Indels** | 64,640,220 | 2,319,524 | 3.59 | 7,512,617 | 11.62 | 32,502,591 | 50.28 | 22,305,488 | 34.51 |

The alternative alleles largely represented the minor allele in the Emirati cohort, i.e., the allele observed in the minority of chromosomes of the cohort; however, they were largely contributed by variants with AF less than 5%, see Figure 1 F). This underscores the applicability and efficacy of the GRCh38 reference genome. When considering polymorphic variants with minor AFs larger than 5%, there are overall 7,693,458 variants in which the alternative allele is the minor allele, i.e. the allele observed in the minority of the Emirati cohort, and 2,320,082 variants in which it is the major allele, i.e. the allele observed in the majority (i.e. > 50%) of the cohort. Thus, for 23.17% of polymorphic Emirati variants, the alternative allele is the major allele, which illustrates the potential and importance of population-specific references.

For the 421,605,069 small variants, we considered all annotated effects, i.e., each variant could have multiple effects on different genes and transcripts, resulting in a total of 756,108,304 molecular effect annotations (see Table 2 and Figure 1, G and H). Most of the variants are unlikely to have a functional effect, since they were intronic (25.28%) or intergenic (25.24%) (Figure 1 H). Most of the variants related to effects that were classified as modifiers, i.e. no specific impact can be directly assigned, are in repeat regions (Table 2; Supplementary Table 5), reflecting repeat region fraction in non-coding genome regions. Notably, about 7.23% of variants have a potential regulatory effect (Figure 1 H). The fraction of variants that alter the gene transcript is 0.283% and thus very low, amounting to 2,138,410 effects, which are typically considered to be of little functional impact. Similarly, there are a few high-impact functional effects on proteins (2,501,565; 0.331%), with only a few of them being common or polymorphic in the Emirati cohort (Figure 1G). Most of these high-impact variants are frameshift variants, of which 422 are polymorphic (AF≥5%) and 2,273 are common (0.01%≤AF<5%) (Figure 1 G, Supplementary Figure 6, Supplementary Table 6).

**Table 2.** Functional profiling of 421,605,069 small variants. This table provides a detailed breakdown of 756,108,304 variant effects on Ensembl transcripts, regulatory regions, and intergenic regions utilizing Variant Effect Predictor (VEP) annotation and arranged based on impact severity from highest to lowest. Effects are stratified into variants in repeat versus non-repeat regions as defined in the UCSC browser for GRCh38. Note that variants can have multiple effects. An exhaustive list of all VEP effects is provided in Supplementary Table 7, which provides additional information about the number and proportions of effects of variants already listed in variant databases.

| Severity | Impact | Effects |  | Effects in Non-Repeat Regions |  |
| --- | --- | --- | --- | --- | --- |
|  |  | [#] | [%] | [#] | [%] |
| HIGH | Frameshift variant | 100,058 | 0.013 | 90,480 | 90.4 |
| HIGH | Stop gained | 67,541 | 0.009 | 64,946 | 96.2 |
| HIGH | Splice donor variant | 53,057 | 0.007 | 44,749 | 84.3 |
| HIGH | Splice acceptor variant | 41,162 | 0.005 | 32,411 | 78.7 |
| HIGH | Start lost | 11,908 | 0.002 | 11,353 | 95.3 |
| HIGH | Stop lost | 6,649 | 0.001 | 5,869 | 88.3 |
| HIGH | Transcript ablation | 40 | 0.000 | 36 | 90.0 |
| MODERATE | Missense variant | 2,145,817 | 0.284 | 2,079,733 | 96.9 |
| MODERATE | Inframe deletion | 41,380 | 0.005 | 34,539 | 83.5 |
| MODERATE | Inframe insertion | 31,242 | 0.004 | 21,756 | 69.6 |
| MODERATE | Protein altering variant | 2,711 | 0.000 | 2,362 | 87.1 |
| LOW | Synonymous variant | 1,082,712 | 0.143 | 1,050,776 | 97.1 |
| LOW | Splice polypyrimidine tract variant | 494,512 | 0.065 | 405,912 | 82.1 |
| LOW | Splice region variant | 436,389 | 0.058 | 368,552 | 84.5 |
| LOW | Splice donor region variant | 87,462 | 0.012 | 76,388 | 87.3 |
| LOW | Splice donor 5th base variant | 33,389 | 0.004 | 28,404 | 85.1 |
| LOW | Stop retained variant | 2,331 | 0.000 | 2,070 | 88.8 |
| LOW | Incomplete terminal codon variant | 1,497 | 0.000 | 1,452 | 97.0 |
| LOW | Start retained variant | 118 | 0.000 | 115 | 97.5 |
| MODIFIER | Intron variant | 191,104,957 | 25.275 | 88,439,212 | 46.3 |
| MODIFIER | Intergenic variant | 190,823,822 | 25.238 | 38,526,270 | 20.2 |
| MODIFIER | Non coding transcript variant | 126,272,380 | 16.700 | 57,321,314 | 45.4 |
| MODIFIER | Downstream gene variant | 59,587,422 | 7.881 | 29,643,722 | 49.7 |
| MODIFIER | Upstream gene variant | 57,520,146 | 7.607 | 28,699,166 | 49.9 |
| MODIFIER | Regulatory region variant | 55,339,899 | 7.319 | 32,599,378 | 58.9 |
| MODIFIER | NMD transcript variant | 49,121,558 | 6.497 | 23,565,443 | 48.0 |
| MODIFIER | Non coding transcript exon variant | 10,403,124 | 1.376 | 7,674,049 | 73.8 |
| MODIFIER | TF binding site variant | 4,920,885 | 0.651 | 3,391,720 | 68.9 |
| MODIFIER | 3 prime UTR variant | 4,918,843 | 0.651 | 3,758,040 | 76.4 |
| MODIFIER | 5 prime UTR variant | 1,402,780 | 0.186 | 1,167,006 | 83.2 |
| MODIFIER | TFBS ablation | 38,811 | 0.005 | 24,196 | 62.3 |
| MODIFIER | Mature miRNA variant | 7,797 | 0.001 | 6,027 | 77.3 |
| MODIFIER | Coding sequence variant | 5,770 | 0.001 | 5,241 | 90.8 |
| MODIFIER | Regulatory region ablation | 135 | 0.000 | 45 | 33.3 |
| **Sum** |  | **756,108,304** |  | **319,142,732** |  |

We observed that 38% of the population’s short variants (143,975,466) were not yet recorded in gnomAD (v4.1), ExAC (v0.3), dbSNP (v154), and ClinVar (as of January 2023) (Supplementary Table 2). A significant subset of these variants (89,964,077; 62%) constituted singleton variants found in only one sample, representing 21% of all identified small variants.

Combined across the 43,608 samples, Manta called 605,452,963 SVs. Per genome, a median of 5,157 deletions (ranging from 2,704 to 8,011), 6,181 insertions (ranging from 3,798 to 9,824), 63 duplications (ranging from 12 to 107) and 2,482 break-end pairs (ranging from 1,602 to 11,400) denoting more complex structural variation were identified (Figure 1 D), Supplementary Tables 1 and 7). When compared to data from the 1KG cohort, these median per-genome counts in EGP are marginally lower; specifically, 1KG reported median values of 5,321 deletions, 6,956 insertions, 395 duplications, and 2,603 break-end pairs. However, the difference is particularly notable for duplications, where the median count in EGP is substantially lower than in the 1KG dataset. By using counts of overlapping SVs identified in the cohort, we generated ideograms of the distribution of SV regions across the autosomal chromosomes (Supplementary Figure 7) and sex chromosomes (Supplementary Figure 8), with the mapped genomic distribution. The CNV caller identified a total of 26,747,919 CNVs, defined as deletion or duplication events with length 1Kb to 5Mb, with a median of 447 deletions per genome (ranging from 271 to 1,933) and 141 amplifications (ranging from 93 to 1,990) (Figure 1 E), Supplementary Table 1 and 8). Comparing these per-genome figures to the 1KG data, the median deletion count in EGP (447) is lower than in 1KG (540), while the median duplication count in EGP (141) is slightly higher than in 1KG (136). The ranges for deletions and duplications per genome are notably wider in the EGP cohort than in the 1KG ranges (1KG deletions: 425-774; 1KG duplications: 111-294). By using counts of overlapping CNVs identified in the cohort, we generated an ideogram of the distribution of common CNV regions across the autosomal chromosomes (Supplementary Figure 9) and sex chromosomes (Supplementary Figure 10). Ideograms show large and complex overlaps between SV and CNV in challenging genomic regions, including sub-telomeric and pericentromeric areas, as well as regions containing large tandem repeats. The location and spread of these structural variants reflect regions of diversity across the EGP cohort.

We analyzed 208,439 genetic variants predicted to have a high impact on canonical transcripts based on annotations from VEP. Most of these were rare or singleton variants, predominantly comprising frameshift and stop-gained mutations. According to ACMG guidelines, most variants across all categories were classified as variants of uncertain significance (VUS), with benign and likely benign variants being comparatively rare. Among heterozygous singleton predicted loss-of-function (pLoF) variants, 19.35% were classified as P/LP. Homozygous singleton pLoF variants annotated as P/LP constituted 6.77% of all homozygous singleton pLoF variants. Notably, a considerable number of frameshift and stop-gained variants were designated as P/LP (Supplementary Table 9).

For each gene, we summarized the total number of predicted pLoF variants and the corresponding counts of heterozygous and homozygous carriers. After filtering, 6,878 genes harbored at least one canonical P/LP pLoF variant (either heterozygous or homozygous), whereas 6,260 genes showed only heterozygous carriers. Genes with six or more homozygous carriers were grouped into a “6+” category (Supplementary Figure 11). Notably, 618 genes had at least one homozygous pLoF carrier within our cohort of 43,608 individuals, indicating that complete loss of function in these genes is apparently tolerated. Such deviations suggest that for these specific genes, the pLoF alleles may be relatively common or under exceptionally low selective pressure, potentially due to biological safeguards such as paralogous compensation or even misclassification of variant pathogenicity, aligning with previous findings.^28,40^

### 3.3 The EGP cohort spans the genetic continuum between European, Asian, and African populations and covers two Arabian components

Unsupervised admixture analysis on the EGP cohort was first conducted to explore latent population structure. We identified an optimal number of five ancestral components (K = 5) using a bootstrapping approach in which 1,000 individuals were randomly sampled from the cohort across 100 replicates. Cross-validation error was minimized at K = 5, indicating this as the most stable and informative level of genetic structure within the EGP population (Supplementary Figure 29). To contextualize the genetic variation observed in the EGP cohort relative to global reference populations, we subsequently performed supervised admixture analysis using the five continental groups from the 1KG population components (European (EUR), South Asian (SAS), American (AMR), African (AFR), East Asian (EAS)) to each sample (Supplementary Table 10; Figure 2 A). Each ancestry component exhibited substantial variability across the cohort, with a range of 0% to 99% (92% for East Asians; Figure 2 F), representing a continuum in ancestry component contributions and indicating considerable heterogeneity in the population. Ranked by average ancestry representation, the EGP components were (± SD) 48% ± 30% European, 25% ± 23% South Asian, 16% ± 21% American, 9% ± 15% African, and 2% ± 5% East Asian. We subsequently stratified the EGP cohort by primary admixture component and identified individuals with a unique ancestry component of more than 90% to support the subsequent population genetic investigations (Figure 2 D).

**Figure 2.**
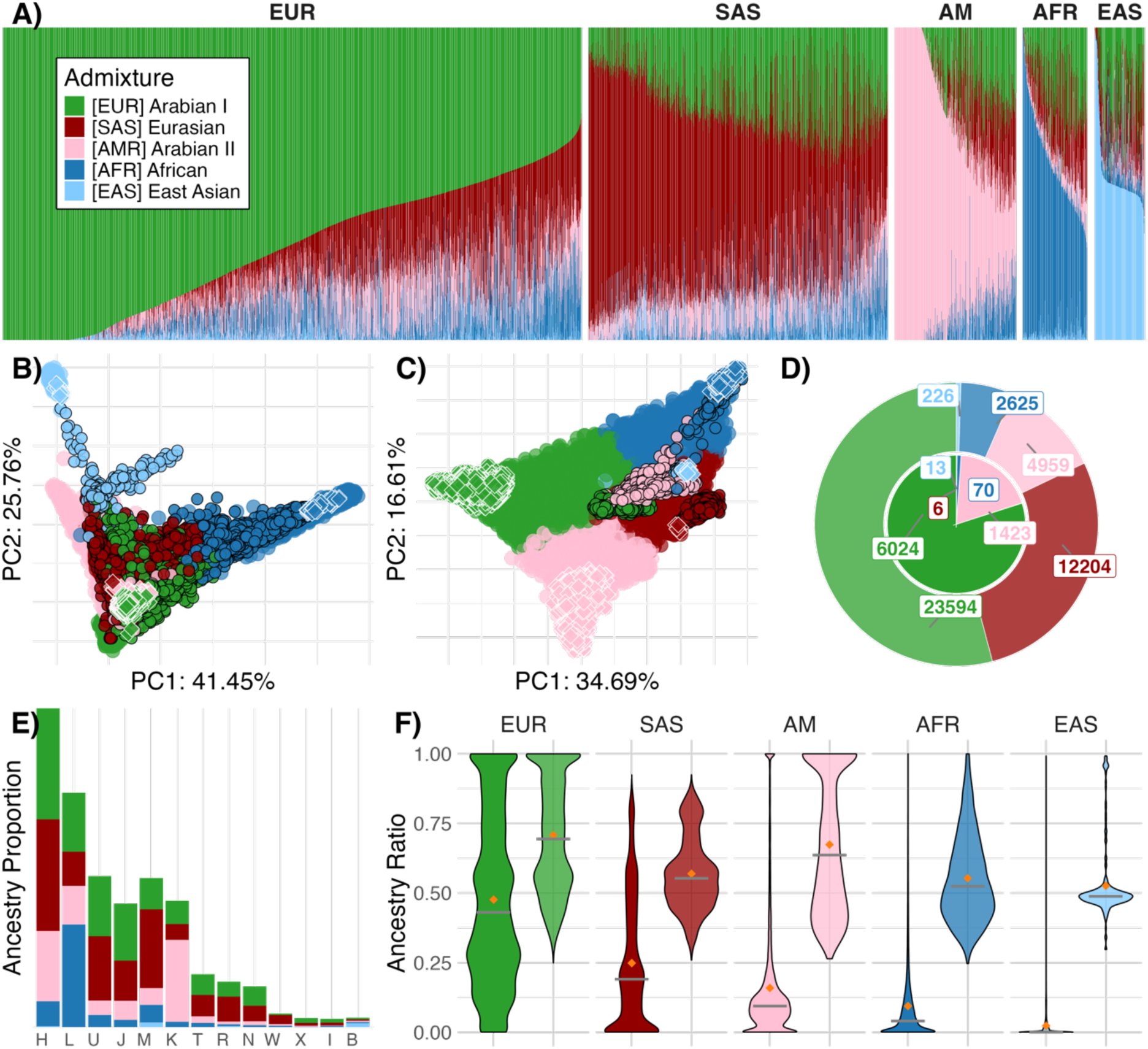
Population stratification of the cohort. This study cohort underwent a supervised admixture analysis utilizing the 1000 Genomes phase 3 (G1K) super populations, e.g., Europe, South Asia, America, Africa, and East Asia, as reference. Every sample was assigned a main ancestry component reflecting the component with the highest value. A) Bar plots show the assignments of ancestry component ratios for the cohort stratified by primary ancestry component contribution to the cohort. Within the sub-panels, the samples were ordered in descending order by the value of the respective main component. B) Principal Component Analysis of the cohort by projection of samples onto the axes defined by the G1K reference dataset. The samples are colored according to their main ancestry component (black frame), and samples with primary component values exceeding 90% are highlighted with diamond shapes (white frame). C) Depicts the projection of G1K samples (black frame) onto the axes defined by the ERGP cohort. Diamond shapes (white frame) show the ERGP samples with primary ancestry components exceeding 90%. D) Pie chart of the primary ancestry component distribution in the cohort. The outer ring shows the distribution over all samples, and the center ring shows the distribution for the subset of samples where the main component exceeds 90%. E) Distribution of main mitochondrial haplotypes stratified by primary ancestry component and sorted by set size. F) Violin plot depicting the value distribution for ancestry components for the full cohort and additionally subsetted for samples with primary ancestry component exceeding 90% (higher opacity).

Based on the directions of maximum global genetic variability, represented by EGP samples projected onto the PCA of the 1KG samples, the EGP population was widely spread across the European, African, and Asian components and covered the region spanned by the European-Asian and European-African axes in proximity to the 1KG European population (Figure 2 B and Supplementary Figure 12 A). When considering the directions of maximum genetic variability within the Emirati population, there was a continuum between three extremes. To investigate the EGP PCA more closely, we colored each individual by its main ancestry component, which showed a consistent picture of continuous transition related to varying ancestry component levels. We further distinguished individuals with more than 90% of the global ancestry components to delineate the major sources of genetic variation in EGP. We find that African and East Asian main ancestry EGP samples overlap with 1KG samples, indicating that these resemble the ancient source of this ancestry component. South Asian 1KG samples and EGP samples, however, are located in close proximity in the PCA plot, with 1KG South Asians likely serving as a proxy for a Western Eurasian or Persian component described earlier.^41^ EGP samples with high European and American 1KG ancestry cluster separately from the respective 1KG populations and are located at the extremes of the EGP-based PCA (Figure 2 C and Supplementary Figure 12 B), thus clearly not representing these 1KG populations, but indeed two components that are not yet represented by 1KG (Figure 2 C) and that align with those discovered by previous genotype PCA for UAE genomes.^42^ In the following, we call these components Arabian I, for which the European component is the closest proxy, and Arabian II, for which the American component is the closest proxy. Consistent with this interpretation is the finding that there is a considerable number of EGP individuals with very high percentages and even exclusively one of these two Arabian components (n=6,024 (13.8%) for Arabian I and n=1,423 (3.3%) for Arabian II, Figure 2D). In contrast, there are almost no individuals having high percentages of the Western Eurasian/Persian, African, or East Asian components. Although the Eurasian component has a considerable average contribution of 25% and is the main component in 28.0% of individuals, we observe only six individuals with more than 90% Eurasian component (Figure 2D and 2F). The assignment of the ancestry components covered by EGP is supported by mitochondrial haplogroups (see Figure 2E). All major global haplogroups are covered by EGP, with a large contribution of haplogroups commonly attributed to Europe (H), the Middle East (J, T, U), Asia (M), and Africa (L). As expected from continuously varying, considerable contributions of often all four major ancestry components (Arabian I, Arabian II, Eurasian, African) across EGP, we find all mitochondrial haplogroups in all major ancestry-stratified groups (Figure 2E). However, when subsetting to individuals with a unique ancestry component of more than 90%, mitochondrial haplogroups are clearly related to ancestry components. We find that J and T are seen in the Arabian I and Arabian II individuals, as are the less prevalent N and X (Supplementary Figure 13). Similarly, L haplogroups 1 to 5 are, with few exceptions, linked to the African main component, R to the Arabian I component, and H to Arabian I and II (Supplementary Figure 14). Interestingly, about 28.8% of individuals (n=1,429) with Arabian II main component had the basic haplogroup K assigned (Supplementary Table 11), with more than 600 individuals with Arabian II component greater than 90% carrying basic K (Supplementary Figure 14). They largely share the same 17 and lack the same seven haplogroup K variants required for a bona fide K haplogroup assignment, which indicates a population bottleneck or genetic isolation. Conversely, haplogroups H, J, and U (covering U1 to U9 branches) are very diverse in EGP, occurring also in their basic form (Supplementary Figure 15), which (as opposed to K and N; Supplementary Figure 16) supports their origin close to or on the Arabian peninsula. ^43,44^

### 3.4 Consanguineous pedigrees characteristic of the UAE are part of the EGP cohort

To characterize pedigrees covered by the cohort, we selected all relationship clusters that covered at least one child and both parents. This stratification resulted in 4,658 individuals grouped into distinct families when considering only the 5,607 parent–offspring relationships (Supplementary Table 12). A total of 264 families (511 individuals) were identified, of which half (264 individuals) were classified as nonconsanguineous and the other half (247 individuals) as consanguineous, i.e., parental relatedness of 3rd or 4th degree.

### 3.5 The EGP uniquely allows investigations related to extensive homozygosity

To explore the patterns of ROH across the EGP cohort, we analyzed the relationship between the number and cumulative length of ROHs for each sample, stratified by main ancestry components (Figure 4A). The median cumulative ROH lengths groups were highest in individuals of Arabian I main ancestry (76,366 kb), followed by Arabian II (75,418 kb), East Asian (72,198 kb), Eurasian (69,031 kb), and African ancestries (66,917 kb) (Supplementary Figure 17A). ROH counts demonstrated substantial variability between groups (Supplementary Figure 17B). The minimum ROH count ranged from 47 in individuals of Arabian I main ancestry to 644 in those of East Asian main ancestry. Maximum ROH counts were highest in the East Asian group (2,381) and lowest in the African group (1,082). Median ROH counts were highest for individuals of Arabian I main ancestry (1,334), followed closely by those of Eurasian (1,276), Arabian II (1,262), and East Asian (1,208) ancestries, while individuals of African ancestry exhibited the lowest median count (745).

**Figure 4.**
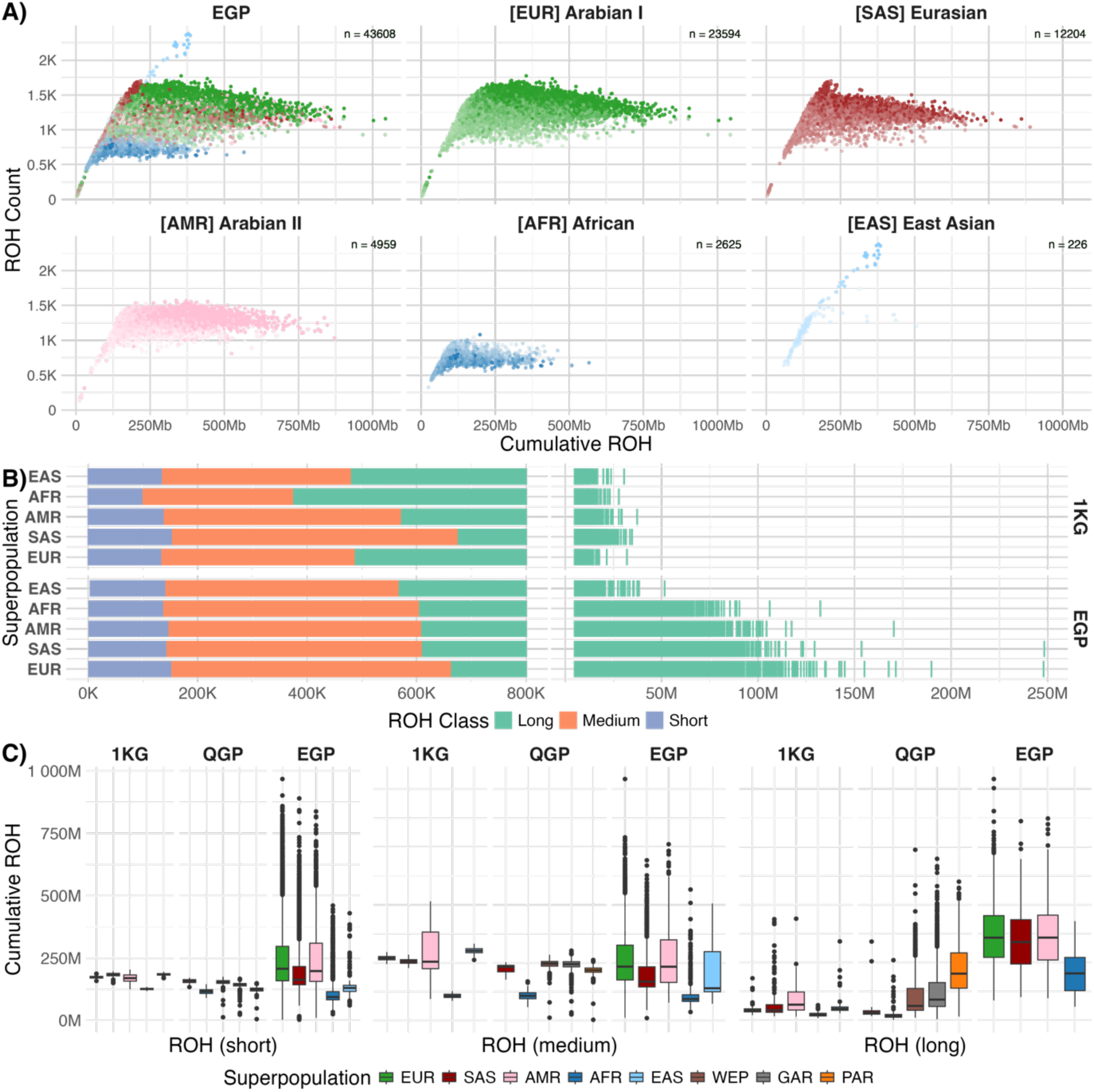
Runs of Homozygosity are stratified by the main ancestry component and subsetted according to classification by size. Subfigure A) shows a scatterplot of the ROHs that ordinate each sample by the number of regions versus the total length of the ROH. The panels show the whole cohort and stratification by main ancestry component, respectively. The color gradient reflects the level of the major components in each sample, i.e., a lighter color is representative of a lower value for the assigned main component. The major component level increased as the number of ROHs increased across all components except for the African main component, where the level decreased as the number of ROHs increased, indicating a unique, inverse relationship between these two variables for this component. The distribution of ROH segments colored by size class is shown in Subfigure B). Similar to the findings reported by Razali et al., a shift towards larger class boundaries can be observed for the long ROH class. Notably, we could not integrate a clean representation of the Qatar reference dataset in this case. In Subfigure C), the panels show ROH stratified by assigned classes (short, medium, long) according to Gaussian admixture clustering as utilized by Razali et al. The samples were stratified within the panels according to the reference dataset and main ancestry component.

A clear ancestry-specific trend was observed on cumulative lengths and counts for ROH (Figure 4A): individuals with higher proportions of Arabian I ancestry, which overlaps with previously characterized Bedouin-related lineages^7^, showed a higher number and longer ROHs, consistent with high consanguinity rates. In contrast, African ancestry was associated with fewer and shorter ROHs.^8^ East Asian ancestry showed high ROH counts, but with lower cumulative length, reflecting reduced consanguinity yet possible population bottlenecks. Our findings are consistent with patterns reported by Mezzavilla et al. (2022) in the Qatari population, where cluster 1 (Bedouin-like) and cluster 3 (Palestinian/South-Asian) showed the highest ROH burdens, while Cluster 2 (African-rich) had the lowest.^45^ However, our East Asian component showed slightly lower ROH loads than their East Asian cluster (Cluster 4), suggesting population-specific variation in ROH architecture that merits further investigation.

Using Gaussian mixture model clustering, ROHs were classified into short (ROH length < 14.73 Mb), medium (ROH length > 14.73 Mb and < 63.03 Mb), and long (ROH length > 63.03 Mb) categories (Figure 4 B and C). The EGP cohort exhibited a shift toward longer ROH, reflecting the high prevalence of consanguineous unions, consistent with findings by Razali et al., 2021.^41^ Stratification by ancestry revealed persistent differences in mean ROH lengths across all categories (Supplementary Figure 17 C). For instance, individuals with African main ancestry consistently exhibited shorter ROHs, likely reflecting greater genetic diversity and lower levels of recent relatedness.

Both frequent and infrequent ROHs show non-random genomic distribution (Supplementary Figure 18; Supplementary Table 13). Infrequent ROHs were enriched near telomeres (Supplementary Figure 19), likely due to local recombination rates. This observation aligns with previous findings and can be attributed to correlations with average recombination rates, as evidenced by earlier research (Supplementary Figures 19-20).^10^ Regions with frequent ROHs were found to have significantly lower recombination rates than infrequent ROHs (p < 0.0001, t-test), suggesting an association between ROH prevalence and genomic recombination dynamics (Supplementary Figures 18-21). Despite these distinct patterns in the distribution and recombination dynamics of ROH regions, enrichment analyses did not reveal significant functional biological pathways in either frequent or infrequent ROH regions.

The inbreeding coefficient (IC), calculated as the cumulative length of ROHs over genome length, was assessed for the EGP cohort and compared to the Qatar Genome Project (QGP) and the 1KG Project reference datasets (Figure 5E and Supplementary Table 14). The Emirati cohort displayed a higher mean and maximum inbreeding coefficient (0.064 and 0.33) than the 1KG Project (0.01 and 0.027) and the QGP cohort (0.048 and 0.248). These findings are consistent with previously reported values of the inbreeding coefficient for the UAE population^46^. When stratified by main ancestry components, individuals with African and East Asian ancestry exhibited the lowest mean inbreeding coefficient and standard deviation, highlighting reduced IC variability in these groups. Notably, the Arabian I and Arabian II ancestry components exhibit higher mean ICs, which are also reflected in the broader distribution of heterozygosity-to-homozygosity ratios within these cohorts (IQR: 0.22 and 0.27, respectively; Supplementary Figure 5D).

**Figure 5.**
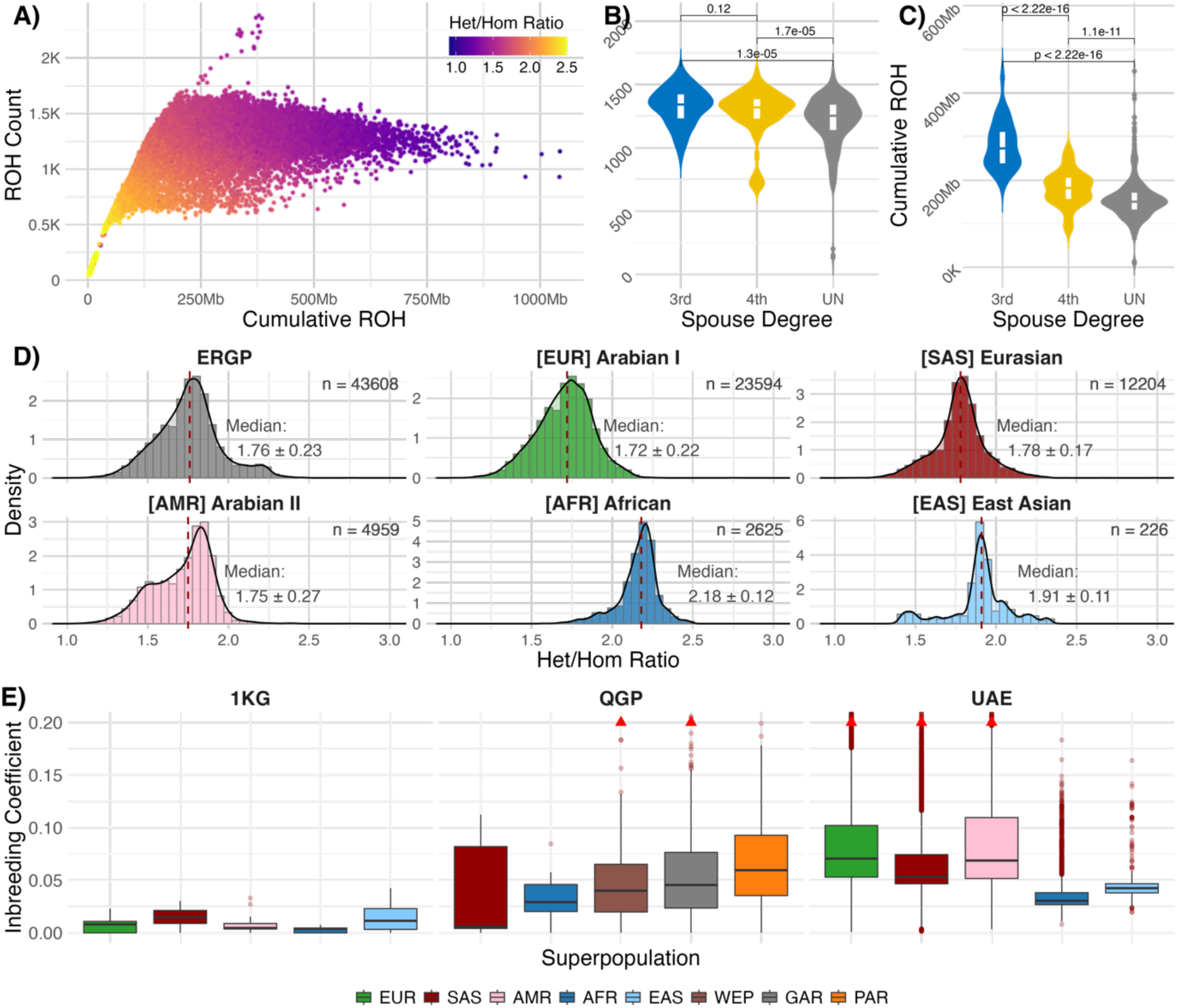
Relatedness and Consanguinity Panel. Subfigure A) shows a scatterplot of the ROHs that ordinate each sample by the number of regions versus the total length of the ROH. The color gradient reflects the heterozygous to homozygous rate, i.e., darker values indicate a higher number of homozygous calls in line with the number and cumulative length of ROH. Subfigures B) and C) depict a cohort of 511 children. Stratification by inferred parental relatedness delineates three groups: nonrelated parents (264), third-degree relationships (60), and fourth-degree relationships (187). The Mann‒Whitney U test revealed a significant disparity in ROH counts between consanguineous and nonconsanguineous offspring, alongside substantial distinctions in ROH lengths across all groups. Subfigure D) depicts the distribution of heterozygous to homozygous rates for the whole cohort and stratification by main ancestry component, respectively. While the median values are comparable to the ones reported by the 1000 Genomes project, the spread reflects the expected high consanguinity, particularly in the [AMR] Arabian II main component. Subfigure E) shows the Inbreeding coefficient as a function of cumulative ROH length over genome length stratified by the main ancestry component. From left to right, the panels show the 1000 Genome reference population, the Qatar reference population, and the Emirati cohort.

The total number and cumulative length of ROHs in offspring were analyzed based on parental relatedness (Supplementary Figures 22 and 23). Stratification delineated three groups: unrelated parents (n=264), offspring of third-degree relationships (n=60), and of fourth-degree relationships (n=187). While the number of ROHs was not significantly different between offspring of third- and fourth-degree consanguineous parents, a significant (p<0.001) shift was observed in the total length of ROHs (Figure 5B and 5C). The density plot (Supplementary Figure 22) and Gaussian clustering analysis (Supplementary Figure 23) of ROH total lengths across the entire cohort demonstrated that offspring from third-degree consanguineous parents presented longer ROH segments. The variability in length, particularly between third-degree and unrelated groups, suggests a similar spread in ROH lengths. Notably, the fourth-degree group appears to be transitional, indicating diverse outcomes in consanguineous relationships, which may not uniformly result in elongated ROH regions. Further stratification by ancestry components (Supplementary Figure 24) revealed nuanced variations in ROH lengths within each relatedness category. Offspring with African and East Asian ancestry exhibited distinct density distributions, suggesting a population-specific influence on ROH patterns. However, the absence of complete family data for certain ancestry components, particularly East Asian, limited interpretability for these groups. Stacked bar plots in Supplementary Figure 23 highlighted the proportional distribution of ROH length categories, demonstrating that third-degree related offspring had the highest proportion of long ROHs, corroborating the strong influence of parental consanguinity on ROH patterns.

### 3.6 The EGP detects variants implicated in disease and affecting molecular function

We investigated the overlap between the identified top 5% frequent and infrequent ROH regions in the cohort and known autosomal dominant (AD) and autosomal recessive (AR) disease genes. Based on ClinGen annotations (Supplementary Figure 25; Supplementary Table 15), 933 AR and 584 AD genes with definitive disease-gene associations were identified on autosomes (chr1-22). Given that the total lengths of the frequent and infrequent ROH regions identified in this study were nearly identical (∼136 MB each), collectively representing approximately 4.7% of the autosomal genome (136 MB / 2904 MB, hg38), the expected number of AR genes within these regions was calculated to be approximately 44 (4.7% of 933), and the expected number of AD genes was approximately 27 (4.7% of 584).

However, analysis of the actual overlap revealed significant deviations from these expectations (Supplementary Table 16). Specifically, we observed an enrichment of AR genes within the frequent ROH regions, while AD genes showed lower-than-expected enrichment within the frequent ROHs. Conversely, AD genes were enriched within the infrequent ROH regions. These distinct patterns of enrichment and depletion suggest differential selective pressures or population histories associated with frequent versus infrequent ROHs and highlight the potential impact of these genomic features on the distribution of disease-associated variants at a population level. We further investigated the prevalence of P/LP Mendelian disease variants in the EGP cohort by summing their AFs per gene (Figure 6 and Supplementary Table 17). The gene with the highest cumulative AF of 4.00%, to which 25 distinct P/LP variants contribute, was *USH2A*. Variants in this gene are reported to cause hearing loss in the UAE^47^ as well as Usher syndrome^48^ and visual impairment^49^ in other Middle Eastern countries. Gene *HBB* has the second-highest cumulative disease variant AF of 2.82%, with 28 distinct variants contributing and causing beta-thalassemia complemented by gene *HBA2* with a cumulative AF of 1.74% of 15 distinct variants causing alpha-thalassemia, both prevalent throughout the Middle Eastern and North African region^50^. P/LP variants in the gene *CBS* cause Homocystinuria due to Cystathionine Beta-Synthase Deficiency, a metabolic disease that, according to the curated database of the Center of Arab Genomic Studies,^51^ has been reported in five MENA countries, but not yet in the UAE. Variants in the gene *BTD* have a cumulative AF of 2.56% for 12 distinct variants in our cohort; however, this is largely driven by the D444H variant, which is common and benign in the homozygous state. D444H may only contribute to partial biotinidase deficiency when present in trans with a more severe mutation, so the clinical impact of *BTD* variants in our cohort should be interpreted in the context of compound heterozygosity^52^. P/LP variants in *ABCA4* cause Stargardt disease 1, a retinal disease, and have a cumulative frequency of 2.48% contributed by 46 distinct variants. The disease has been described in five MENA countries, including the UAE^53^. In addition, gene-based cumulative P/LP variant AFs highlight a significant burden in other genes associated with hearing and vision loss, such as *GJB2*, *SLC26A4*, and *EYS*. *CYP1B1*, a well-known gene linked to primary congenital glaucoma, emphasizes the significant impact of eye-related disorders, while *PAH*, associated with phenylketonuria, points to metabolic disorders. *PKHD1* highlights the burden of renal diseases, particularly autosomal recessive polycystic kidney disease (ARPKD). Furthermore, genes such as *MPL* (hematological disorders) and *MEFV* (familial Mediterranean fever) highlight a diversity of conditions with relevance to the Arab population.^54,55^ Of the genes with the highest P/LP variant AF, most follow an autosomal recessive inheritance pattern, with reported cases often observed in offspring of consanguineous parents.^56–58^ However, consistent with recessive inheritance patterns observed for the top genes, none of them is located in the 5% most infrequent ROH regions, but also only two of them are located in the 5% most frequent ROH regions, *PKHD1* and *EYS*.

**Figure 6.**
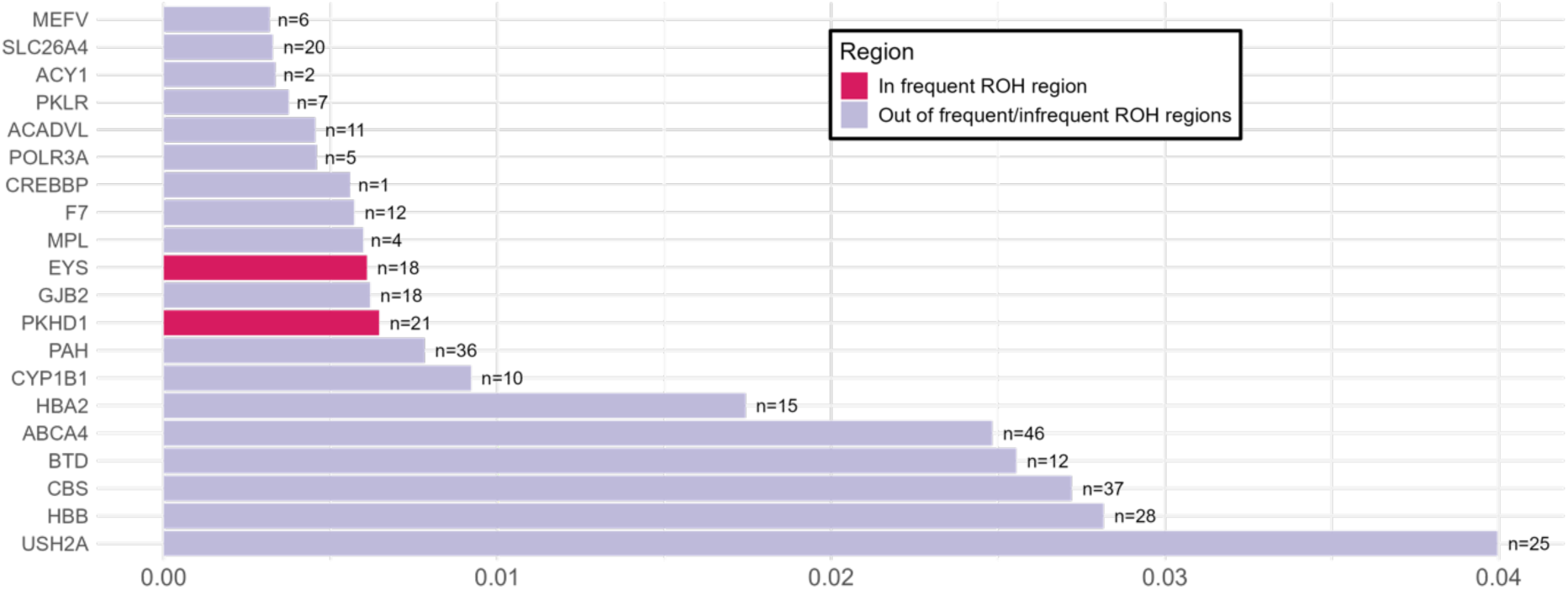
Cumulative Allele Frequency of Pathogenic/Likely Pathogenic Variants in Mendelian Inherited Genes. This plot displays the top 20 Mendelian inherited genes with the highest cumulative allele frequency (AF) of pathogenic (P) and likely pathogenic (LP) variants in the UAE cohort. The x-axis represents the cumulative AF of P/LP variants in the UAE cohort, with the number of observed P/LP variants (n) indicated for each gene. Genes located in frequent runs of homozygosity (ROH) regions are highlighted in red. None of the top 20 genes shown here are located in infrequent ROH regions. The full list of genes and their corresponding data are listed in Supplementary Table 16.

When investigating AFs cumulated for Mendelian diseases, we find that most P/LP variants are located in genes attributed to the aforementioned diseases, i.e., thalassemia and related diseases, Usher syndrome, hearing loss, and retinal diseases (Supplementary Figure 26, Supplementary Tables 18 and 19). Finally, when combining P/LP variant AFs across disease categories, about 1,000 distinct genetic variants relate to severe recessive diseases, congenital structural abnormalities, inborn errors of metabolism, and intellectual disability, respectively. This is followed by retinal disorders (n=450 P/LP variants) and hearing loss (n=250 P/LP variants). This highlights the wealth of information covered by EGP that can be utilized for improving clinical diagnostics of rare diseases in the future.

We further investigated population-specific variants. Supplementary Table 20 details the annotated variants (n = 206) with frequencies exceeding 2% in the Emirati population and those that are rare (frequency < 0.1%) in public databases, such as gnomAD and Bravo. Most of the variants are of VEP impact ‘MODIFIER’, indicating that their functional impact is unclear. Five variants that are common in EGP have a predicted deleterious effect on genes *ADAM29*, *HOXD12*, *LACTBL1*, *PCDHAC1*, and *SOS1*, respectively (VEP impact ‘HIGH’). Other variants in these genes are associated with a range of different, important phenotypes such as blood pressure, BMI, schizophrenia and cognitive function measurements, but association studies in Emirati cohorts will be needed to elucidate associations and specific functional effects of the predicted deleterious effects that are to date reported with high frequencies only in EGP.

Additionally, high-impact prevalent variants, frequent, and infrequent ROH regions are listed in Supplementary Table 16. Among them are the previously mentioned variants, which are interestingly predominantly found in ROH regions (*ADAM29* (91.55% in ROH), *HOXD12* (100%), *LACTBL1* (84.37%), *PCDHAC1* (98.18%), *SOS1* (100%)) and may in the case that the variant effect is as deleterious as predicted be considered as gene knockouts.

### 3.7 Ploidy estimation indicates differences in sexual development and highlights large aberrations affecting chromosomes 19 and 22

A total of 141 individuals were affected by sexual or autosomal disorders, denoted by deviations from expected median chromosomal coverages (Supplementary Figure 27). Of these, 93 individuals, constituting 0.21% of the cohort, exhibited sex chromosome aneuploidies, including Klinefelter syndrome (XXY) identified in 41 individuals, Turner syndrome (XO) identified in 23 individuals, XYY syndrome identified in 16 individuals, and triple X syndrome (XXX) identified in 12 individuals (Supplementary Figure 28). In one case, exact sex chromosome ploidy could not be determined reliably, but was most compatible with XXY (Klinefelter syndrome).

Among the autosomal aneuploidy disorders, Down syndrome (trisomy 21) was identified in 41 individuals. Furthermore, our analysis revealed partial chromosome 22 and 19 gains close to a full additional chromosome in two and four samples, respectively, that may be due to mosaicism. Trisomy of chromosomes 22 and 19 can have severe consequences, including lethality, in cases of full trisomy of the entire chromosomes. Therefore, the six collective samples presented with mosaicism in these chromosomes may mitigate the clinical impact.

## 4. Discussion

In this manuscript, we present a comprehensive characterization of the genetic landscape of the UAE population, drawing insights from the genomes of 43,608 individuals, which contain a total of 421,605,069 variants. We have developed a variome catalogue, which includes alleles and AFs, as well as the effects of variants on gene expression, protein function, and pathogenicity, and integrated this information into a user-friendly web interface for querying this database.

We observed that a considerable proportion of detected variants had not been recorded in variant databases yet. The singleton category is most affected by random sequencing errors, where one false positive in one sample is enough to be counted as a singleton variant. Therefore, singletons should be interpreted carefully. The observed singleton percentage in our cohort (30.86%) is comparable to that observed in the Qatari (26%) and Russian (33%) cohorts^59,60^. However, other Asian variomes showed higher percentages (45%-55%) for singletons^61–64^, which may reflect differences in variant calling, processing, and filtering procedures. Our study revealed that the frequency of rare variants (those with a prevalence of less than 0.1% but not a singleton) was 58.83%. This aligns closely with the frequency observed in the gnomAD^65^ database, where 60% of the variants have an AF <0.1%.

The genetic makeup of the EGP cohort displays ancestral heterogeneity, with significant contributions from at least two Arabian, an Eurasian, and an African component. Meanwhile, African and East Asian genetic components, though present, exerted a smaller influence on the overall genetic composition. Notably, three of the 1KG Project ancestry components were identified, with the help of unique ancestry individuals and their mitochondrial haplogroups, to represent proxy components for two indigenous Arabian components and a Eurasian component. This reconciles the results obtained in this study, which employed a global and supervised approach, with the results of non-supervised approaches that identified the same components for Qatar^41^ and the UAE^42^. Future in-depth population genetic characterization will further characterize these and potential sub-components on a local geographic scale in addition to the global perspective provided in this study.

Since we have no phenotypic data available for the present study, we cannot assess the accuracy of P/LP variants classified as pathogenic or likely pathogenic according to ACMG, and the reported frequencies should be considered an upper bound. Still, this study highlights that given population-based information about P/LP variation needs to be followed by clinical investigations of patients to develop targeted genetic screening and counseling programs, particularly for the Arab population in the UAE, to reduce the burden of Mendelian disorders and enable early interventions. The clustering of sensory, metabolic, and hematological disorders reflects unique regional challenges, underscoring the need for comprehensive public health initiatives.

Overall, the diversity observed within the Emirati population’s genetic makeup is comparable to that of neighboring populations, including those of Qatar^41^ and Kuwait^66^. This genetic heterogeneity is attributed to the prehistorical migration patterns within the Arabian Peninsula and the influence of the endogamous culture^42^. The EGP cohort shows a strong influence of an Arabian component (46%), with a considerable subset of individuals exclusively exhibiting this component. This identified ancestry components roughly align with those of previous genome-wide^42^ and immunogenetic ancestral haplotype studies^67^, which indicated a more dominant influence of South Asian over European components within the UAE population. Mitochondrial haplogroups identified in EGP also relate to those identified in a cohort of Emirati females, in which 15 different haplogroups were detected that are typically attributed to Africa, East Asia, and the Near East^68^. In the current study, the larger sample size and wider range of variants enabled a more comprehensive and accurate representation of the population’s complex genetic landscape, which may not have been fully captured in earlier reports. Previous research, including both whole-genome and mitochondrial genome analyses, demonstrated that there is no clear association between birthplace and genetic stratification within the contemporary population of the UAE^2^. This observation suggests that the population’s genetic makeup was established well before the delineation of modern political borders, with the split times of the two Arabian components estimated to be 12,000 years ago and of the Arabian component from the Eurasian component 20,000 years ago, based on data from Qatar^41^.

One of the most striking observations from the large EGP cohort is the continuous spectrum of genetic variation and contributions from diverse ancestry components. Although we stratified individuals by their predominant ancestry components in this study, such discrete grouping is not ideal for downstream applications. This is because individuals typically carry substantial contributions from multiple components, resulting in a gradient of admixture proportions across the cohort. A more refined approach may involve stratifying individuals based on their ancestry at specific genomic regions rather than relying solely on global ancestry estimates.

Due to the elevated consanguinity rates in the region, we observed an expected increase in the total length of ROH within the cohort. This was especially demonstrated when (1) comparing offspring of third- and fourth-degree consanguineous parents (p<0.0001) and (2) when analyzing ancestral components, with the Arabian I ancestral component exhibiting the highest median number of ROHs and the African ancestral component showing the lowest number of ROHs. Overall, the findings suggest varying levels of homozygosity based on the ancestral component. We identify chromosomal aneuploidies reliably and with expected prevalences. In addition to well-described syndromes affecting the sex chromosomes and chromosome 21, we identify a few cases with chromosome 19 and 22 gains that, to the best of our knowledge, have not been described so far and which seem to be related to mosaicism.

Our findings demonstrate that, although the EGP cohort comprises heterogeneous ancestral groups, there has been an increase in genetic drift within the population and genetic isolation from other populations. This is likely due to indigenous Arabian components that are partly affected by genetic isolation and recent consanguineous parental relationships. This is demonstrated by population-specific variants, especially in ROH patterns within the population. Finally, applying the EGP variome to future projects for subjects originating from the Middle East will aid in further refining and elucidating Mendelian disease variants detected in EGP.

## Future Perspective

Insights from the allele frequency distribution and the discovery of variants not yet reported in variant databases underscore the need for population-specific reference genomes. This is particularly critical in investigating variants of uncertain significance (VUS) and interpretation in a clinical setting due to the undefined impact on disease risk or phenotype and lack of sufficient evidence, as evidenced by databases such as ClinVar. This challenge is further amplified for ancestral groups that are significantly underrepresented in genomic databases. Recent research indicates that individuals who are not of European ancestry exhibit a considerably higher VUS identification rate than their European counterparts^69^. Data from the Middle East were not included in this report, revealing the deficiency of representation of this ancestral group in important analyses. The EGP enriches global scientific efforts by providing a comprehensive dataset that adds to the diversity of available genome data.

By offering a detailed view of the genetic landscape of the population, the EGP empowers policymakers to draft targeted healthcare strategies and make informed decisions tailored to the unique genetic characteristics and needs of the population. For example, variants reported to cause Mendelian disease, as well as high-frequency reference variants, can be investigated specifically for the Emirati population, improving the diagnostics of common and rare monogenic disorders, while allelic and variant frequencies could advance pharmacogenomic applications.

We propose that these benefits can be captured by employing several key measures: (i) refining the accuracy of the current disease-causing gene panel used in screening, (ii) potentially broadening the spectrum of tested variants, (iii) identifying genetic components related to disorders that are comparatively rare worldwide but show a higher prevalence in the UAE, and (iv) improving the integration of pharmacogenomics in the healthcare system of the UAE^70,71^.

An important future direction for genomic research, especially within genetically diverse populations, involves implementing a pangenome strategy to establish a more comprehensive reference genome. The traditional variome reference genome, while valuable, represents a limited, flat perspective, often based on a narrower genetic sampling that may not capture the full spectrum of human genetic diversity observed in contemporary populations. This limitation is especially pronounced in heterogeneous populations, such as the population of the Middle East, where the genetic variance is vast, and the representation in global reference genomes is insufficient. The pangenome approach will address these challenges by providing a reference that encompasses the genetic diversity of a population. In parallel, future studies should also focus on systematically comparing allele frequencies of fine-mapped, causative variants associated with common diseases—particularly those identified through co-localization and functional studies—to better understand population-specific genomic risk profiles. Therefore, for the UAE population, which is known for its rich genetic heterogeneity, adopting a pangenome strategy would be a significant step forward in population genetics and genomic medicine.

## Author Contributions

RMO, MM, IW, NAM, and HA were involved in the conceptualization of this project. TM, JM, and JQ were involved in laboratory operations, sequencing, and the development of DRAGEN-output files. RMO, MM, IW, AAA, AA, NAM, and HA were involved in the design of the methodology, visualization, and data analysis. RMO and MM were involved in the preparation of the initial draft, with input from IW, AAA, AA, NAM, HAN, TM, JM, JQ, MA, MSM, ND, YI, RH, GT, SI, and HA. All authors have critically reviewed the work. This study was supervised by FA, AAM, and HA. All authors agreed to the final version of the manuscript.

## Ethics Statement

The study was approved by the Department of Health – Abu Dhabi (DOH: DOH/CVDC/2022/1701) and the Khalifa University Research Ethics Committee (H21-045). All procedures were conducted in accordance with the ethical guidelines and regulations of these committees and in accordance with the principles outlined in the Declaration of Helsinki. Written informed consent was obtained from all participants before their inclusion in the study.

## Data Availability

Data access for this study is provided through the Emirati Reference Genome Program (EGP) website hosted via the Department of Health (DoH). Researchers can request data access by completing the Data Access Agreement available on the EGP website [https://www.doh.gov.ae/en/research/the-emirati-reference-genome-programme]. Upon submission of the signed agreement, the requested data will be made available via a secure Virtual Private Network (VPN). The Data Access Agreement ensures compliance with all relevant ethical, legal, and security protocols, outlining the terms for data usage, confidentiality, and the purpose of the research. Additionally, the MAF table is available at the European Genome-phenome Archive (EGA) under accession ID EGAD50000001558. Access to the EGA-hosted dataset requires submission of a Data Access Committee (DAC) application in accordance with EGA’s controlled-access protocols. All data access and handling procedures are designed to maintain the integrity and confidentiality of the dataset, in line with applicable data protection regulations. The data processing and analysis code is accessible on CodeOcean with a provisional DOI: 10.24433/CO.6975178.v1.

## 1.0 Supplementary Text

### 1.1 Sample Collection

Participants who agreed to the study’s objectives, as detailed in the participation information sheet, signed an electronic consent form. The UAE Emirates ID (EID) reader associated the consent with the participant’s personally identifiable information (PII), and an M42 field barcode was issued along with a collection tube at the time of sample collection. To ensure data security, the consent form, UAE EID, contact details, and other pertinent information were encrypted once the record was submitted, with only designated personnel accessing the PII records. The Longenesis Consent blockchain consent management tool (Nurse Application) facilitated the synchronization of registration data. This approach allowed linking the M42 field barcode with the M42 OMICs Laboratory Information Management System (LIMS) to auto-generate an order. These stringent ethical practices ensured the confidentiality, privacy, and voluntary participation of all individuals contributing to this valuable genomic resource.

For consenting participants, 5 ml of blood was collected. The collected samples were delivered to the M42 OMICs lab and associated with the M42 field barcode-generated order. Then, an EGP lab barcode was issued for further tracking and identification. The blood fractionation process involved using the Hamilton easyBlood Star for auto fractionation, ensuring precision and efficiency. The meticulous sample management approach facilitated seamless tracking and ensured the integrity and quality of the collected biological specimens.

### 1.2 DNA Extraction and Sequencing

The sequencing process was carried out using the NovaSeq 6000 S4 Reagent Kit v1.5 (300 cycles) on the NovaSeq 6000 System, with Binary Call (BCL) files generated and subsequently demultiplexed using bcl2fastq on the DRAGEN platform by Illumina. All samples were sequenced to achieve 90 gigabases (Gb) of raw data. In good-quality whole-genome sequences (WGS) and following a high mapping rate, 90 Gb will translate into approximately 30X genome mapping effective coverage, which is the target coverage of the EGP. We allowed some tolerance and accepted WGS with a yield of>87 Gb. Samples below this threshold receive a top-up as described in supplementary section 1.1.

In the Illumina top-up procedure, samples below the 30X autosomal sequencing coverage threshold underwent an additional library preparation using the Illumina DNA PCR-free Library Prep kit on the Tecan Freedom Evo 200 NGS Platform. Barcoding was repeated with the IDT® for Illumina® UMI DNA/RNA UD Indexes Set A, followed by library QC using the Qubit ssDNA Assay kit on the Fluoroskan Plate Reader. The top-up samples were then pooled, with the number of samples depending on the required yield and the capacity of the Novaseq 6000 S4 flowcell. Pooling QC was conducted using the Qubit ssDNA Assay kit on the Qubit 4.0, followed by sequencing using the NovaSeq 6000 S4 Reagent Kit v1.5 (300 cycles) on the NovaSeq 6000 System.

## 2.0 Supplementary Table

**Supplementary Table 1. Quality control (QC) metrics of Illumina samples.** The table demonstrates the QC metrics of 43,608 Illumina samples, including the coverage metrics, mapping and aligning metrics, Q30, and variant calling metrics.

**Supplementary Table 2: Tabular breakdown of SNV and Indel counts across chromosomes: known, novel, and singleton novel variants**. This table summarizes the distribution of genetic variants across the chromosomes, including known, novel, and singleton novel variants. Novel variants are defined as those absent from public databases, as described in the methods. Singleton novel variants are a subset of novel variants that appear only once in the dataset. Major Alternate Alleles (MAA) represent alternative alleles with an allele frequency (AF) exceeding 50% in the population. All percentages are calculated relative to the total number of variants per chromosome, while the percentage of novel MAA is calculated relative to the total MAA.

**Supplementary Table 3. Tabular breakdown of SNV counts across chromosomes.** The table delineates variant categories, encompassing common, polymorph, rare, and singletons. Common variants are observed in more than 0.1% and less than 5% of samples, while rare variants are present in at least two individuals but less than 0.1%. Polymorph variants are observed in more than 5% of samples. Singletons, on the other hand, occur in only a single individual. Accompanied by respective percentages, this data offers detailed insights into variant distribution and prevalence within distinct chromosomes.

**Supplementary Table 4. Tabular breakdown of Indel variant counts across chromosomes.** The table delineates variant categories, encompassing common, polymorph, rare, and singletons. Common variants are observed in more than 0.1% and less than 5% of samples, while rare variants are present in at least two individuals but less than 0.1%. Polymorph variants are observed in more than 5% of samples. Singletons, on the other hand, occur in only a single individual. Accompanied by respective percentages, this data offers detailed insights into variant distribution and prevalence within distinct chromosomes.

**Supplementary Table 5. Tabular summary of all annotated variants.** Functional characterization of variants. The table shows the number of variants classified by their functional impact as annotated by VEP, ordered by impact from most severe to least severe. Additional columns list the respective number of variants inside repeat regions and a sub-classification for the number of novel variants.

**Supplementary Table 6. List of Canonical HIGH Impact Variants.** Tabular breakdown of variants annotated with HIGH impact label by VEP, stratified by variant prevalence, i.e., polymorphic, common, rare, and singleton.

**Supplementary Table 7. Tabular breakdown of Structural Variants (SV) counts across chromosomes.** The table delineates the total structural variants over all 43,608 samples called by Manta in the Dragen pipeline. The present variant types include insertions (INS), deletions (DEL), duplications (DUP), and breakends (BND). The table depicts the counts for the whole genome, the autosomes, and the allosomes. Notably, these counts represent the total number of SVs in all samples without merging, i.e., the variants were counted in each sample.

**Supplementary Table 8. Tabular breakdown of Copy Number Variations (CNV) counts across the cohort.** The table delineates the total CNVs over all 43,608 samples called by the Dragen pipeline. The variant types include deletions (DEL) and duplications (DUP).

**Supplementary Table 9. Tabular Breakdown of pLoF Variants in homozygous state.** Distribution of predicted loss-of-function (pLoF) variants across genes in the analyzed cohort, summarized by the total number of pLoF variants per gene, alongside counts of heterozygous and homozygous carriers. Genes are classified based on the burden of homozygous carriers, with those having six or more homozygous carriers grouped into a "6+" category. This classification facilitates identifying genes with elevated homozygous carrier frequencies, potentially highlighting candidate genes of clinical or functional significance due to increased homozygous loss-of-function burden within the cohort.

**Supplementary Table 10. Tabular breakdown of supervised admixture components.** The table illustrates the average percentage for each of the five admixture components assigned using supervised analysis on 43,608 UAE nationals. Additional median and standard deviation columns show the heterogeneous distribution of components within the cohort.

**Supplementary Table 11.** For each EGP individual the mitochondrial haplogroup and the main ancestry component.

**Supplementary Table 12. Tabular breakdown inferred relationships.** The table illustrates the number and degrees of relationships identified in the dataset. Relatedness within the dataset was determined using the KING relatedness inference algorithm., utilizing a subset of 150,000 variants.

**Supplementary Table 13. List of Frequent and Infrequent Runs of Homozygosity (ROH).** The table lists the Runs of Homozygosity (ROH) illustrated in Supplementary Figure 13. The empirical determination of cutoffs for frequently observed ROH (found in more than 8029 samples) and infrequently observed ROH (found in less than 1500 samples) is detailed in Supplementary Figure 12. A window size of 10 kb was used to calculate the median number of samples in adjacent regions. Centromeric regions were excluded from the analysis. Genes with autosomal recessive (AR) and autosomal dominant (AD) inheritance overlapping these regions are listed in the respective columns. AR and AD genes with a definitive association with diseases were identified using the ClinGen database (downloaded on 2024-12-05). The total length of all frequent ROH regions was 136.02 MB, compared to 135.92 MB for the infrequent ROHs. The frequent ROH regions exhibited a higher overlap with AR genes and a lower overlap with AD genes compared to the infrequent ROH regions. Frequent ROHs overlapped 65 AR genes and 18 AD genes, whereas infrequent ROHs overlapped 38 AR genes and 42 AD genes. In total, the analysis includes 584 AD genes and 933 AR genes. Supplementary Table 16 provides a complete list of this analysis’s AD and AR genes.

**Supplementary Table 14. Inbreeding coefficients (IC) for three cohorts**—EGP (Emirates Genome Project), QGP (Qatar Genome Project), and G1K (1000 Genomes Project)—stratified by five super populations: African (AFR), American (AMR), East Asian (EAS), European (EUR), and South Asian (SAS). Mean and median IC values are reported, highlighting population-specific differences in genetic relatedness and consanguinity. Additionally, unique subpopulations within QGP, such as Gulf Arabian (GAR), Persian (PAR), and Western European (WEP), are included to provide detailed comparisons.

**Supplementary Table 15. List of Autosomal Dominant and Autosomal Recessive Genes**. The table lists genes with autosomal dominant and autosomal recessive inheritance patterns classified as having a definitive gene-disease relationship based on the ClinGen database as of December 5, 2024. Only genes located on autosomes and associated with definitive disease classifications are included.

**Supplementary Table 16. Tabular breakdown of annotated variants prevalent in the UAE population.** The table presents variants commonplace in the UAE population, each exhibiting a frequency surpassing two percent, considering the population’s significant admixture. These variants are notably rare in public databases, such as gnomAD-listed populations and Bravo, with frequencies lower than 0.1%. Variants lacking gnomAD entries were conservatively assigned a frequency of zero and are included in the table, ordered based on their prevalence.

**Supplementary Table 17. Cumulative Allele Frequency of Pathogenic/Likely Pathogenic Variants in Mendelian Inherited Genes.** This table presents the cumulative allele frequency (AF) of pathogenic (P) and likely pathogenic (LP) variants in a curated set of Mendelian inherited genes, as reported by Aamer et al. (2024). The analysis was limited to genes with a "Definitive" gene disease category in ClinGen. The number of P/LP variants observed in the UAE cohort was counted for each gene, and their cumulative AF was calculated. Additionally, the table indicates the location of each gene relative to ROH regions.

**Supplementary Table 18. Cumulative Allele Frequency of Pathogenic/Likely Pathogenic Variants in Mendelian Inherited Diseases**. This table presents the cumulative allele frequency (AF) of pathogenic (P) and likely pathogenic (LP) variants in a curated set of Mendelian inherited diseases, as reported by Aamer et al. (2024). The analysis was limited to diseases linked to genes with a "Definitive" gene-disease category in ClinGen. The number of P/LP variants observed in the UAE cohort was counted for each disease, and their cumulative AF was calculated.

**Supplementary Table 19. Cumulative Allele Frequency of Pathogenic/Likely Pathogenic Variants in Mendelian Inherited Disease Categories**. This table presents the cumulative allele frequency (AF) of pathogenic (P) and likely pathogenic (LP) variants in a curated set of Mendelian inherited disease categories (n=19), as reported by Aamer et al. (2024). The analysis was limited to diseases linked to genes with a "Definitive" gene-disease category in ClinGen. The number of P/LP variants observed in the UAE cohort was counted for each disease category, and their cumulative AF was calculated.

**Supplementary Table 20. Tabular summary of high-impact variants located within ROH regions.** Functional characterization of variants classified as high function impact, as annotated by VEP. Additional columns show the counts of homozygous alternative alleles as well. As the number of samples for whom these variants fall within ROH segments.

**Supplementary Table 21.** Tools and versions used in the respective analysis steps.

## 3.0 Supplementary Figures

**Supplementary Figure 1.**
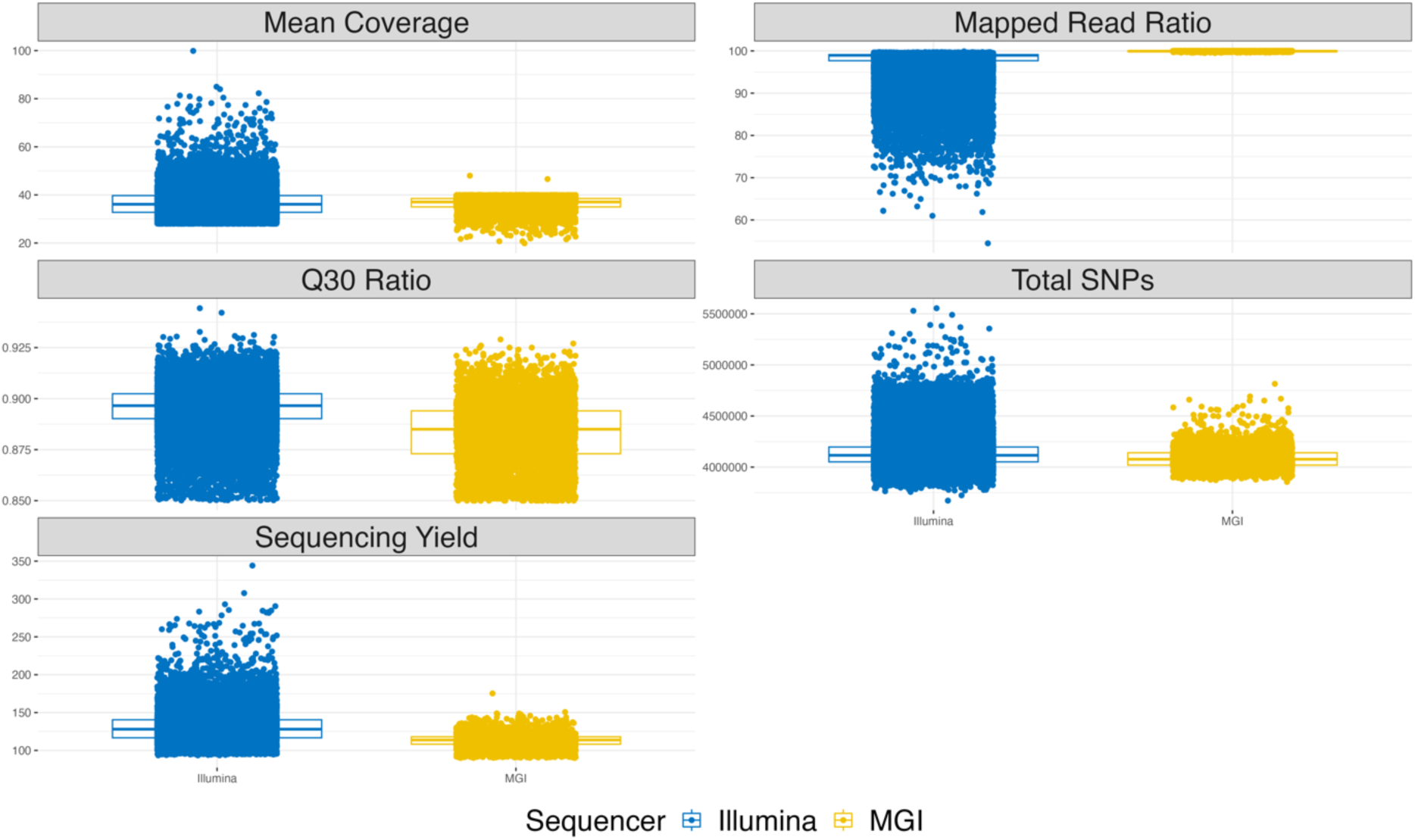
Boxplot Panel Comparing Sequencing Quality Metrics Stratified by Sequencing Technologies: This visual representation showcases five key sequencing quality metrics—Mean coverage, Ratio of mapped reads, Q30 Ratio, Total SNPs, and Sequencing yield—displayed as boxplots. The data is stratified based on the two sequencing technologies utilized: Illumina short read and MGI. This comparative analysis provides insights into the performance and variation observed across these critical quality metrics within the context of the large-scale sequencing project.

**Supplementary Figure 2.**
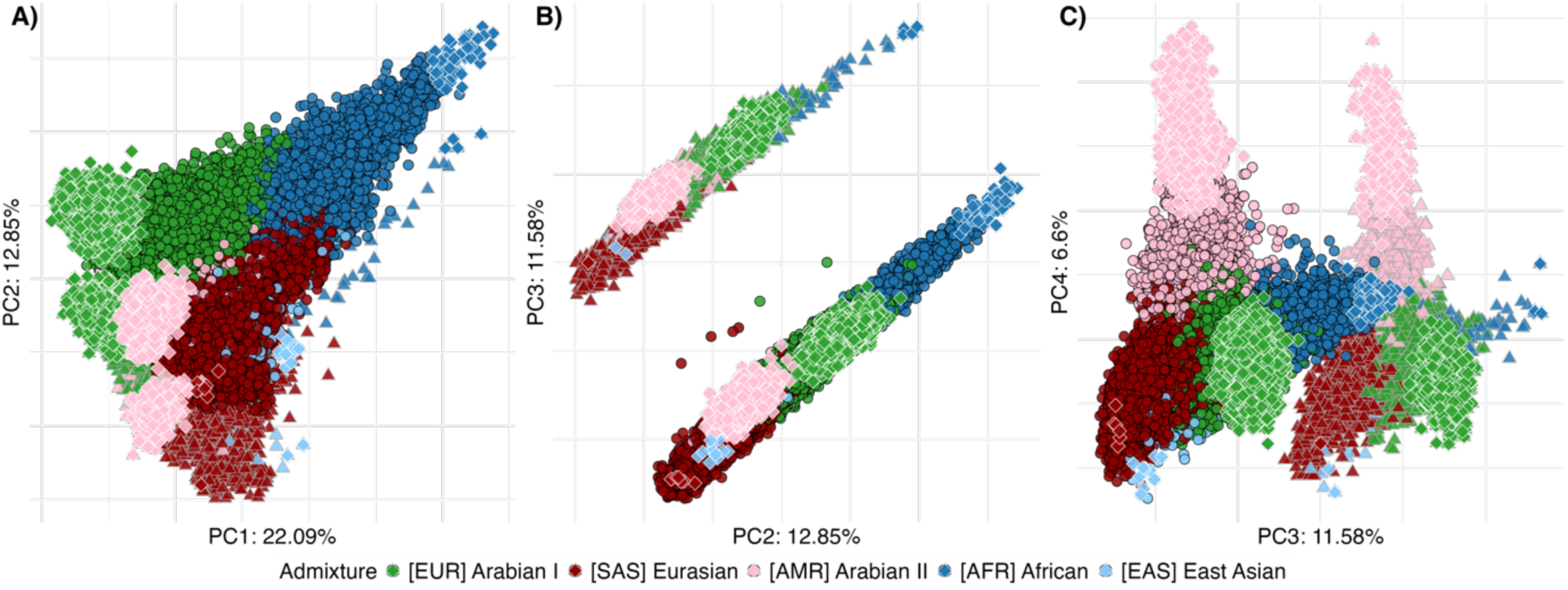
Principal component analysis of overall 50,000 sequenced samples using population-specific SNPs. The principal component ordination plot presents the directions of maximum genetic variation of the cohort of 50,000 individuals, of which 43,608 were sequenced with Illumina and 6,392 were sequenced with MGI. Here, 146,874 population-enriched single nucleotide polymorphisms (SNPs) were used, selected to be protein-coding, and have an allele frequency between 45 and 55%. Each sample is color-coded by its primary admixture component. Panels A) and B) illustrate discernible batch effects between samples sequenced using Illumina and MGI technologies. While both subsets display a consistent ordination pattern reflecting major admixture components, Panel B) notably highlights the distinct separation attributed to the sequencing platforms, especially captured by PC3, but to a lesser extent also by PCs 1 and 2. Panel C) showcases the outcomes of a secondary Principal Component Analysis (PCA) after removing MGI-sequenced samples from the dataset.

**Supplementary Figure 3.**
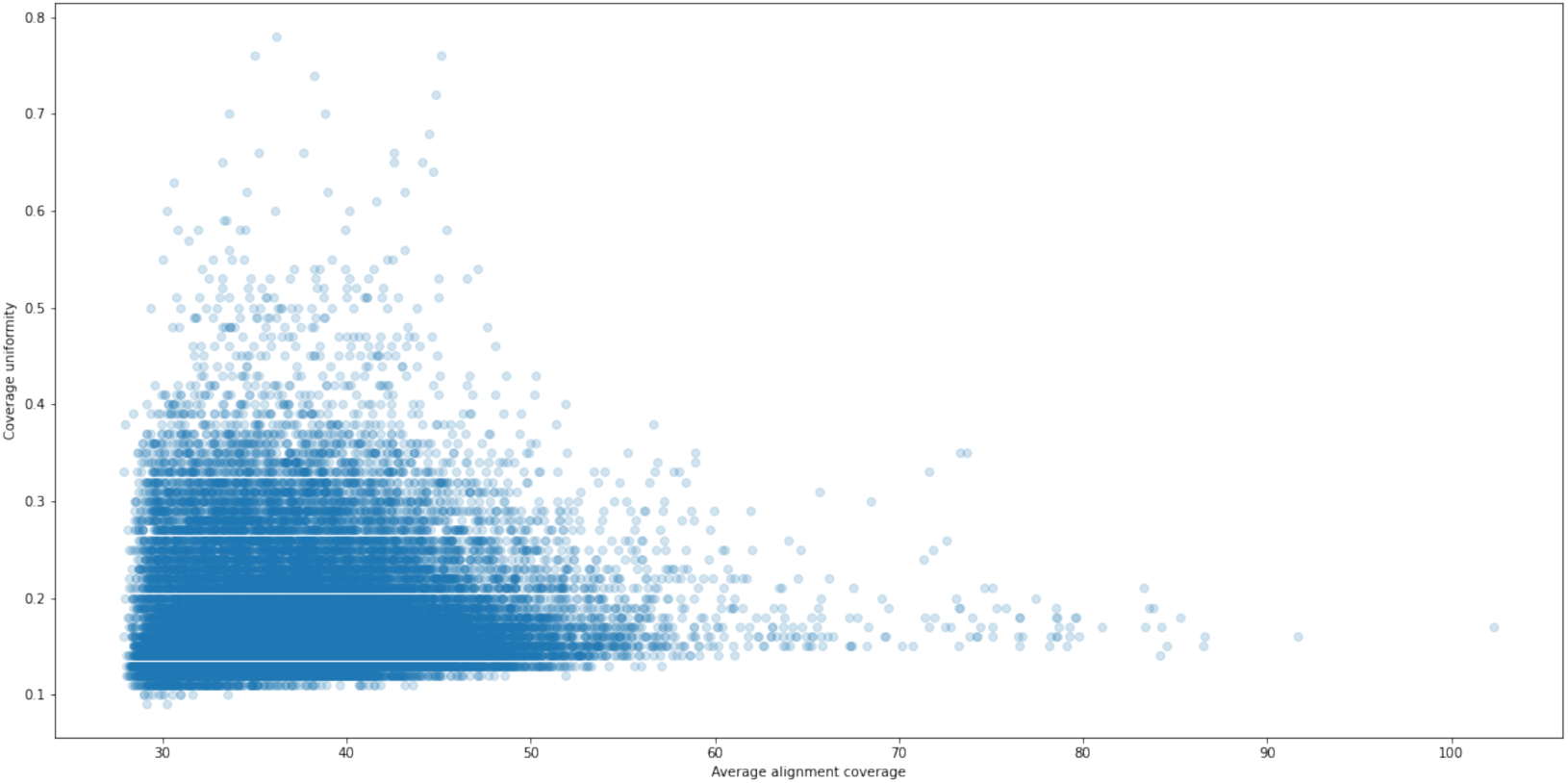
Scatterplot Analysis of Metric Coverage Uniformity Relative to Average Alignment Coverage by DRAGEN. This figure represents the metric coverage uniformity compared to the average alignment coverage, as determined by the DRAGEN (Dynamic Read Analysis for GENomics) platform. Each data point on the scatterplot corresponds to a specific sample, capturing the interplay between two critical metrics. The x-axis denotes the average alignment coverage, reflecting how sequencing reads align to the reference genome. On the y-axis, the metric coverage uniformity is illustrated, representing coverage consistency across the genomic regions under investigation. A closer examination of the scatterplot reveals patterns and trends in the data, offering insights into the performance and efficiency of the DRAGEN platform. The dispersion and clustering of data points provide valuable information about the platform’s ability to maintain uniform coverage across different genomic regions.

**Supplementary Figure 4.**
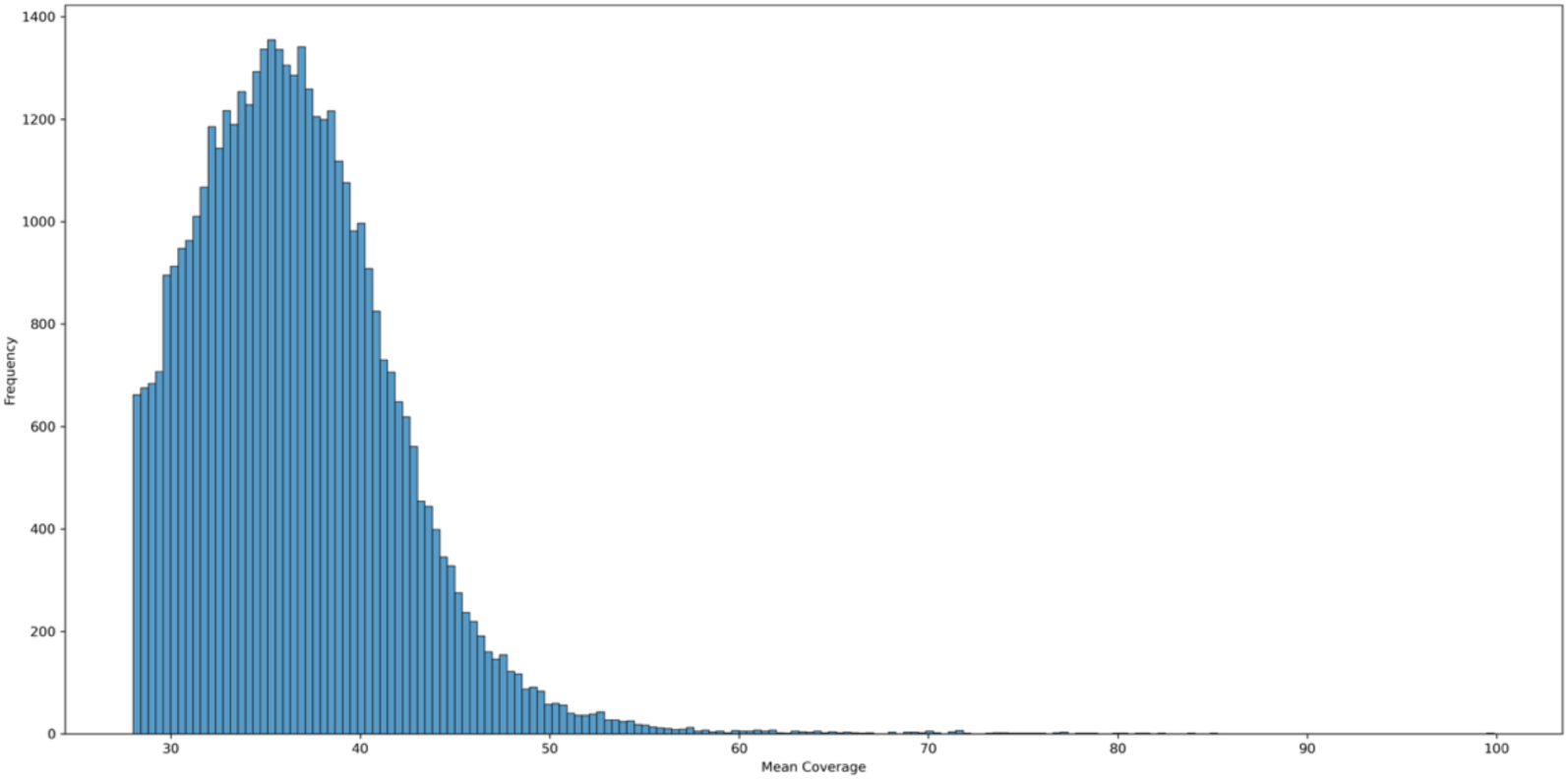
Distribution Analysis of Mean Coverage across Autosomes and Sex Chromosomes. This figure presents a histogram depicting the mean coverage per sample. The x-axis of the histogram represents the mean coverage values, while the y-axis illustrates the frequency or count of samples falling within each coverage range.

**Supplementary Figure 5.**
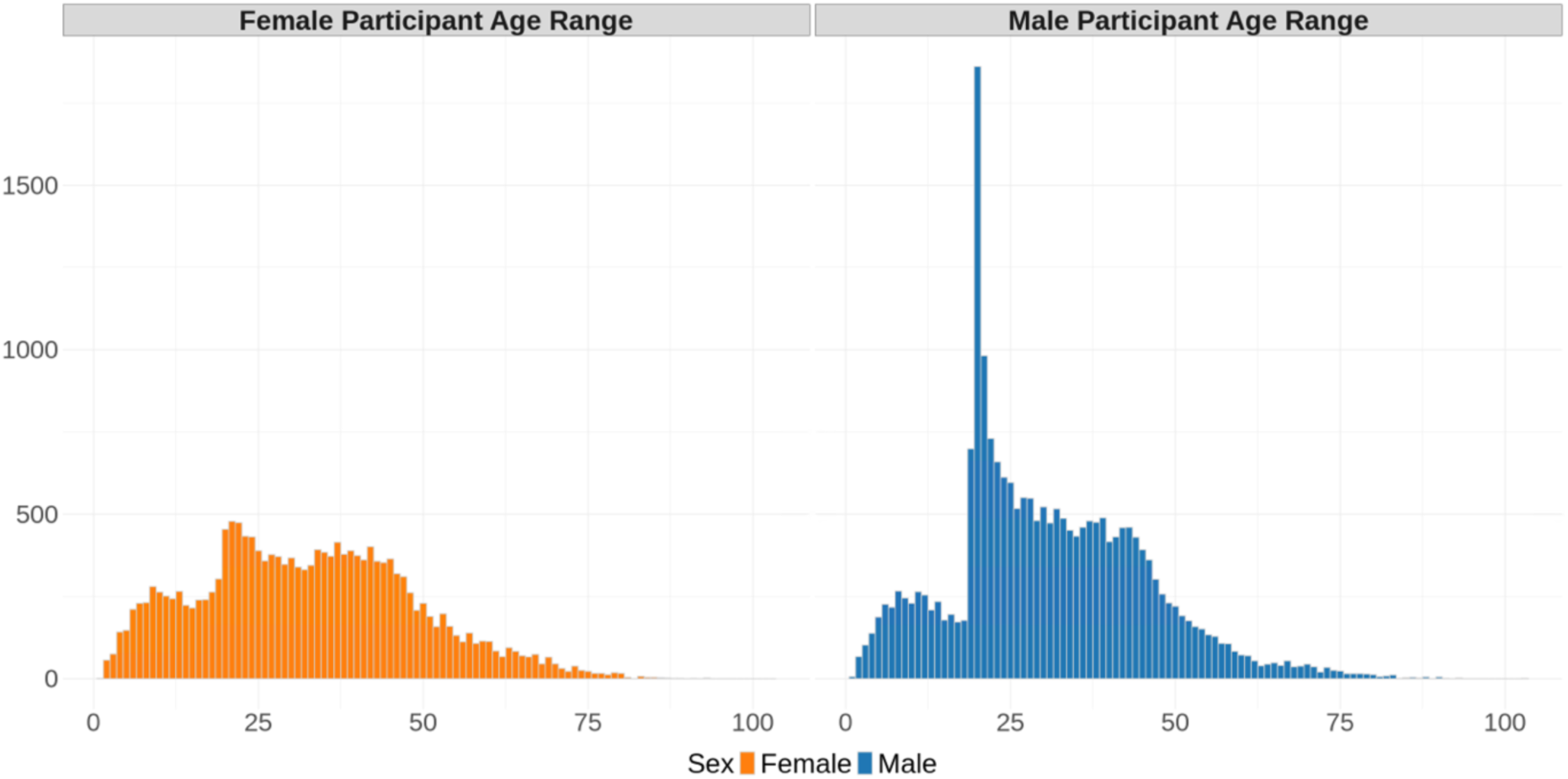
Histograms depict the distribution of participant ages categorized by sex. The left panel illustrates the age distribution of female participants, while the right panel displays the age distribution of male participants. A notable spike in male participants around twenty suggests the influence of sample collection during military service.

**Supplementary Figure 6.**
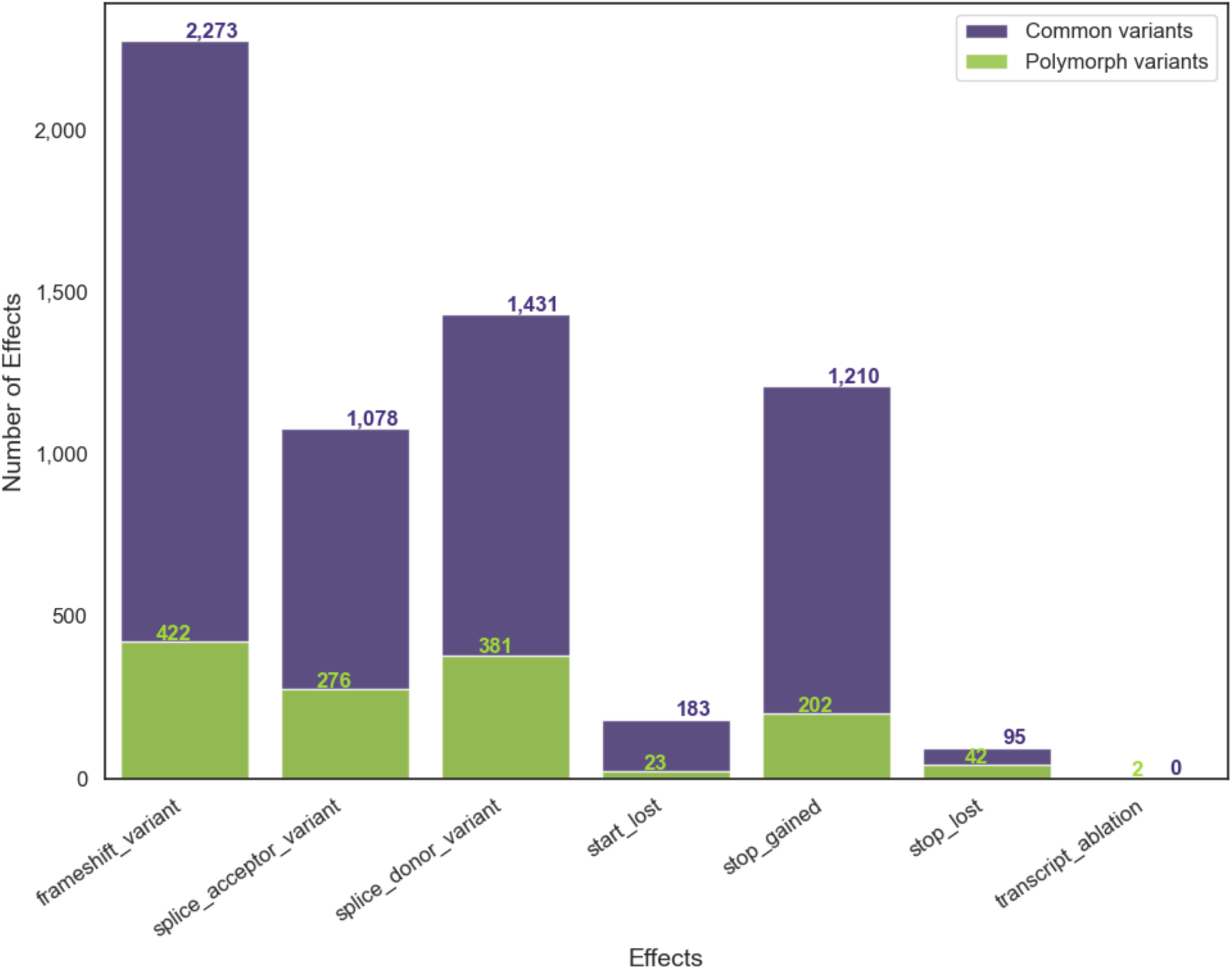
Distribution of common and polymorphic variants stratified by impact category as per VEP annotation. The figure shows the number of effects for the high-impact and canonical genetic variants across the Polymorphic and Common categories for various variant effect types, such as frameshift, stop_gained, etc. These variants were filtered using bcftools to include only high-impact and canonical variants. Common variants are observed in more than 0.1% and less than 5% of samples, and polymorphic variants are observed in more than 5% of samples.

**Supplementary Figure 7.**
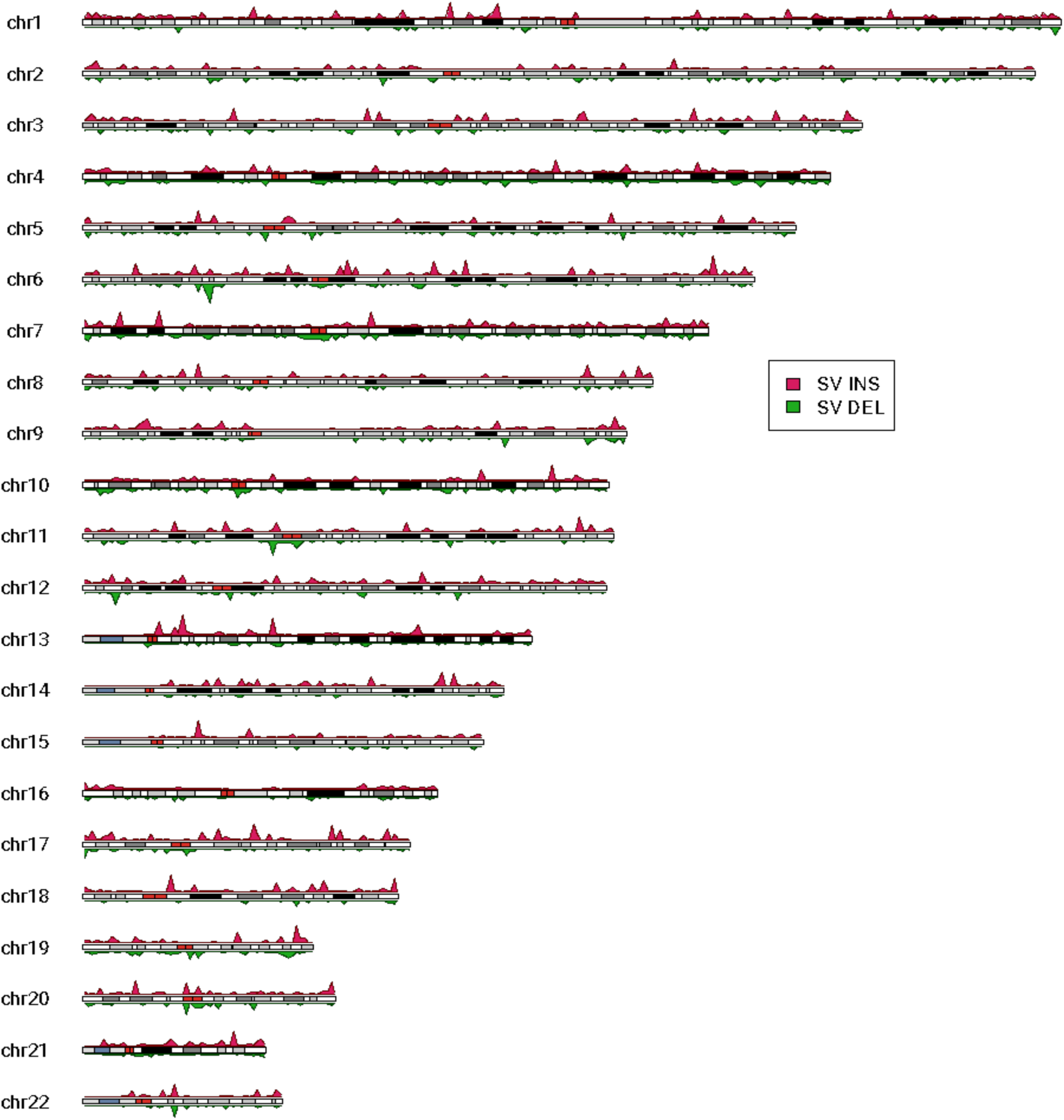
Ideogram Illustrating the Distribution of Structural Variants Regions in Autosomal Chromosomes. This figure presents a combined ideogram and histogram approach to elucidate the distribution of structural variants (SVs) within autosomal chromosomes, observed in a minimum of 4 of 43,611 individuals after filtering to account for the potential error rate of single variant caller manta. The red upper segment represents insertions, while the green lower segment identifies deletions. Utilizing kernel density estimation (KDE), this plot provides a quantitative perspective on SV distribution. The red color within the chromosome represents the centromere.

**Supplementary Figure 8.**
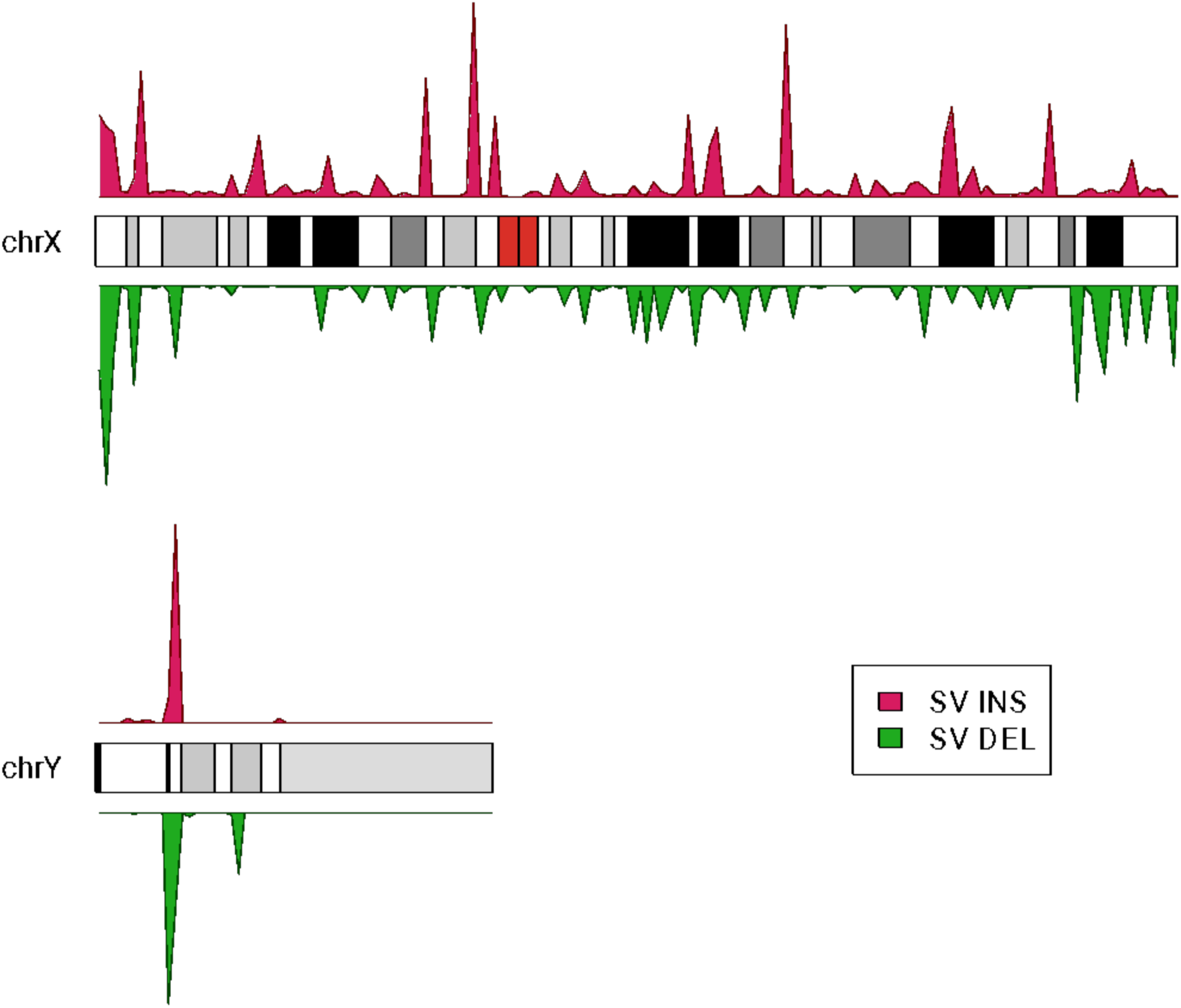
Ideogram Representation of Structural Variant (SV) Distribution in Sex Chromosomes. This karyotype plot visually portrays the distribution of structural variants (SVs) within the sex chromosomes, specifically distinguishing between insertions and deletions. The karyotype is organized into two distinct segments: the upper part, highlighted in red, illustrates insertions, while the lower part, marked in green, represents deletions. The red insertions at the top of the karyotype signify genomic regions where additional genetic material has been incorporated. At the same time, the green deletions at the bottom denote regions where genetic material has been removed. This clear separation visually assesses these SV types’ prevalence and spatial distribution across the sex chromosomes.

**Supplementary Figure 9.**
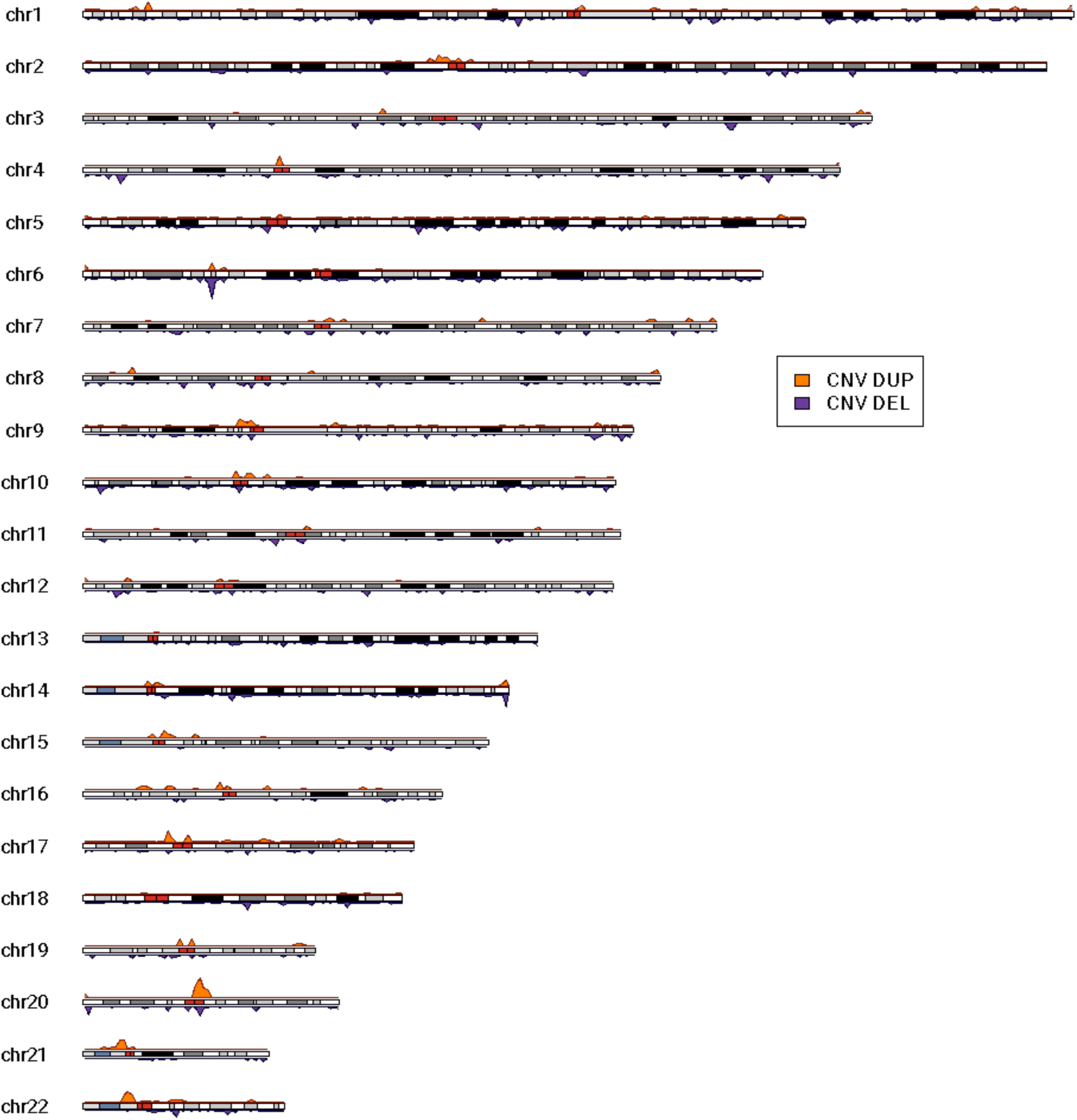
Ideogram Illustrating the Distribution of Copy Number Variant Regions in Autosomal Chromosomes. This figure presents the density distribution of copy number variants (CNVs) within autosomal chromosomes, distinguishing between duplications (depicted in orange at the top) and deletions (illustrated in purple at the bottom). Utilizing kernel density estimation (KDE), this plot provides a quantitative perspective on CNV distribution. The red color in the chromosome represents the centromere.

**Supplementary Figure 10.**
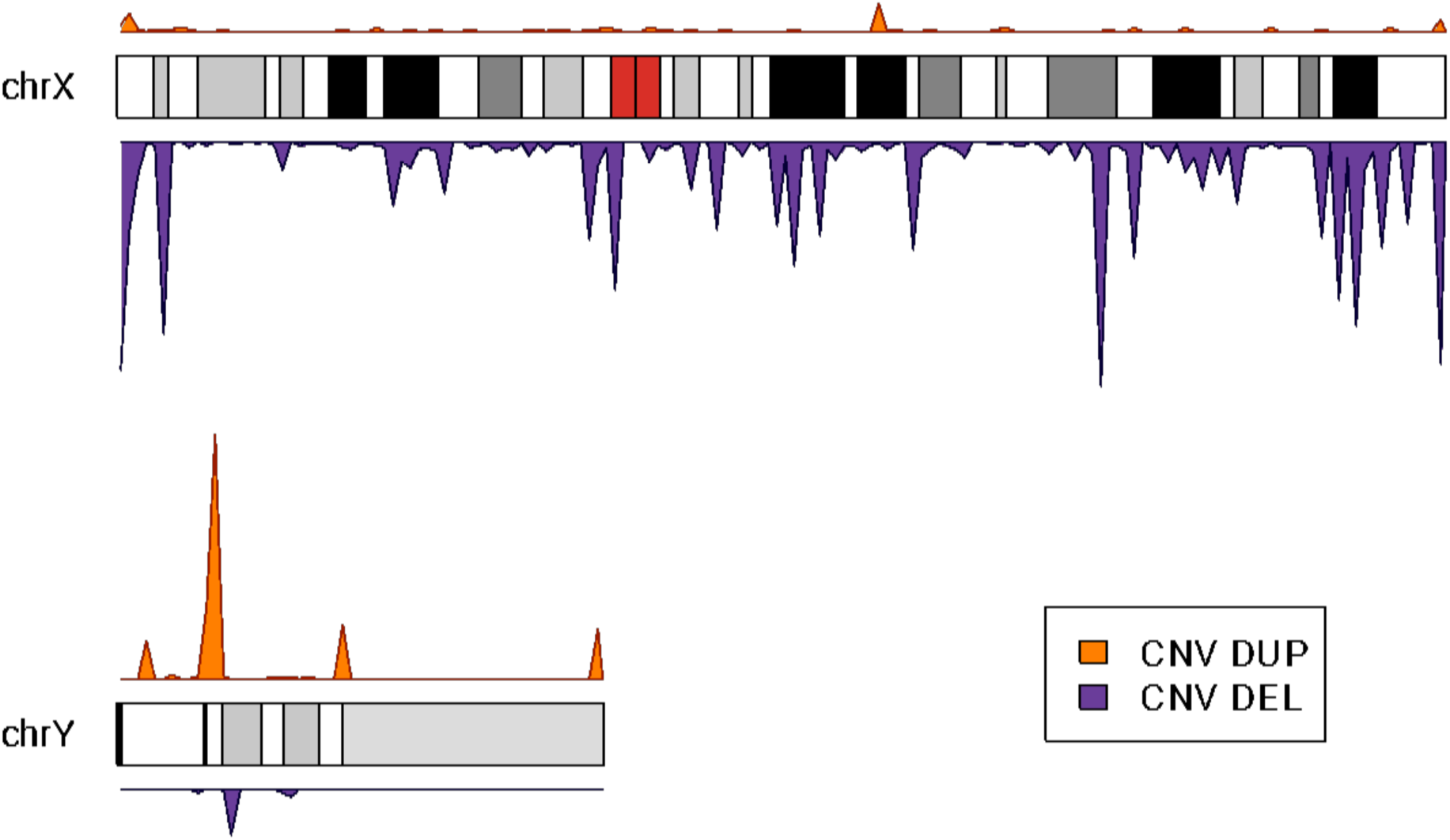
Ideogram Visualization of Copy Number Variation (CNV) Distribution in Sex Chromosomes. This karyotype plot illustrates the distribution of copy number variations (CNVs) within the sex chromosomes, emphasizing the distinction between duplications and deletions. The karyotype is partitioned into two distinctive sections: the upper part, highlighted in orange, signifies duplications, while the lower part, adorned in purple, represents deletions. The orange-colored duplications at the top of the karyotype indicate genomic regions with an increased copy of genetic material. At the same time, the purple deletions at the bottom denote regions where a portion of genetic material has been removed. This visual separation facilitates a quick and comprehensive assessment of the prevalence and spatial distribution of these CNV types across the sex chromosomes.

**Supplementary Figure 11.**
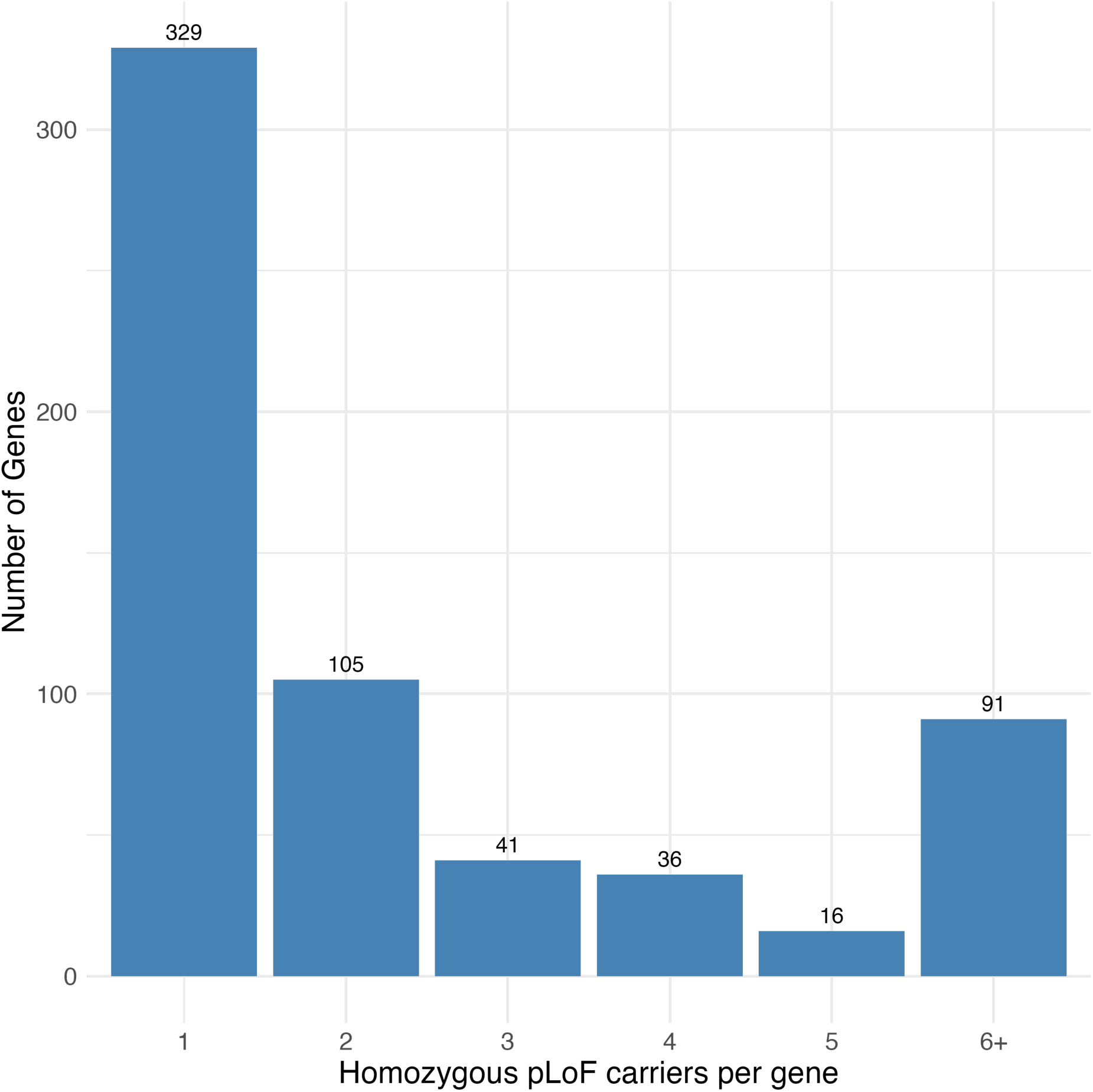
Distribution of genes by number of homozygous LP/P pLoF variant carriers. The bar plot shows the number of genes stratified by the number of individuals homozygous for pathogenic or likely pathogenic (P/LP) predicted loss-of-function (pLoF) variants. Each bar corresponds to a gene count based on the number of homozygous carriers, from one to five, with genes having six or more homozygous carriers grouped into the “6+” category. Gene-level carrier burden was determined by aggregating homozygous carrier counts per gene across the full cohort, using rare (MAF <1%) high-impact pLoF variants annotated on canonical Ensembl transcripts and classified as P/LP according to ACMG criteria.

**Supplementary Figure 12.**
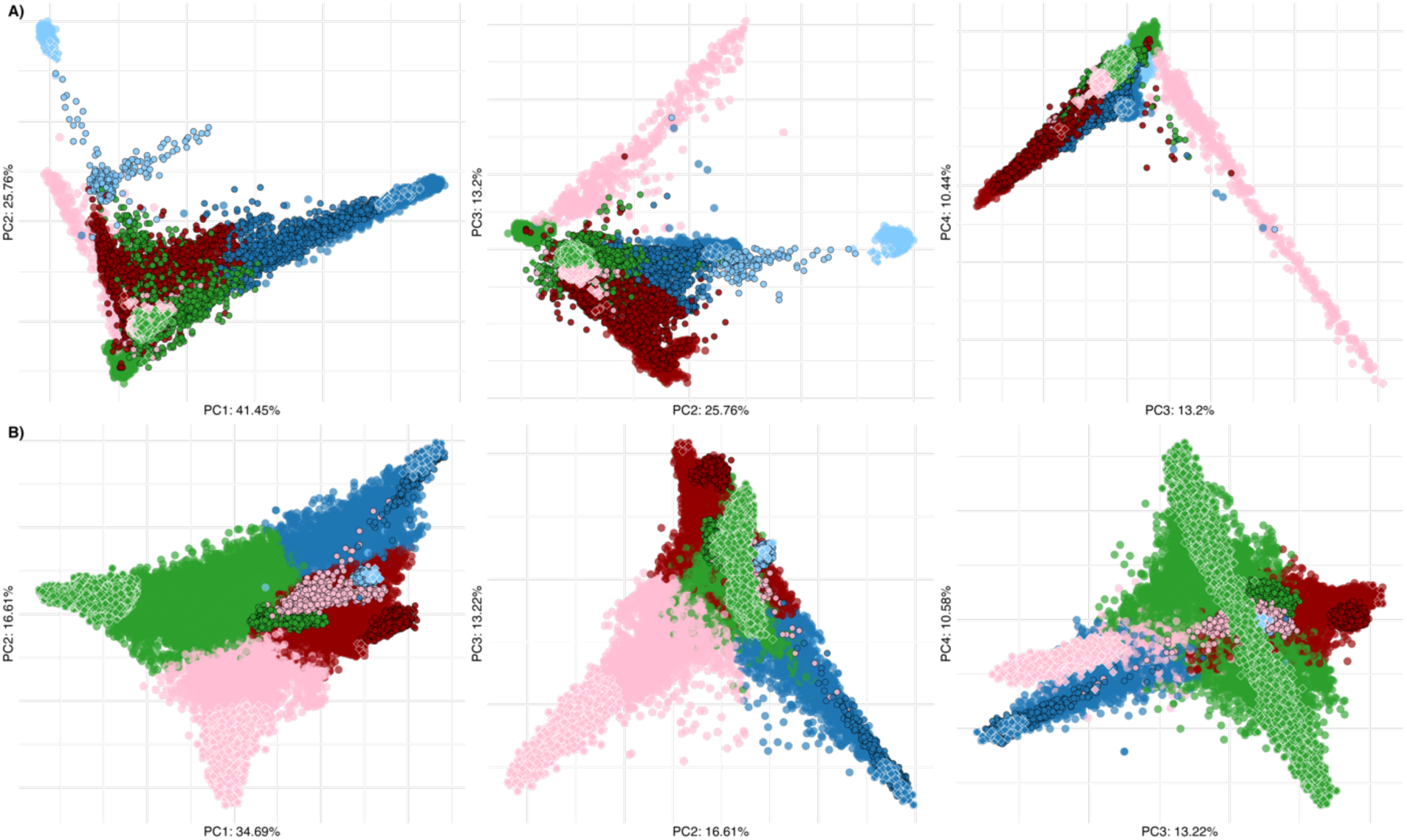
PCA Comparison of UAE National Cohort and 1000 Genomes Reference Samples. This figure presents the principal component analysis (PCA) results comparing the genetic variation among 43,608 UAE nationals to samples from the 1000 Genomes reference dataset. Each panel displays a scatterplot representing the distribution of samples along the first four principal component axes. Subfigure A) shows the Principal Component Analysis of the cohort by projection of samples onto the axes defined by the G1K reference dataset. The samples are colored according to their main ancestry component (black frame), and samples with main component values exceeding 90% are highlighted with diamond shapes (white frame). B) Depicts the projection of G1K samples (black frame) onto the axes defined by the ERGP cohort. Diamond shapes (white frame) show the ERGP samples with main ancestry components exceeding 90%.

**Supplementary Figure 13.**
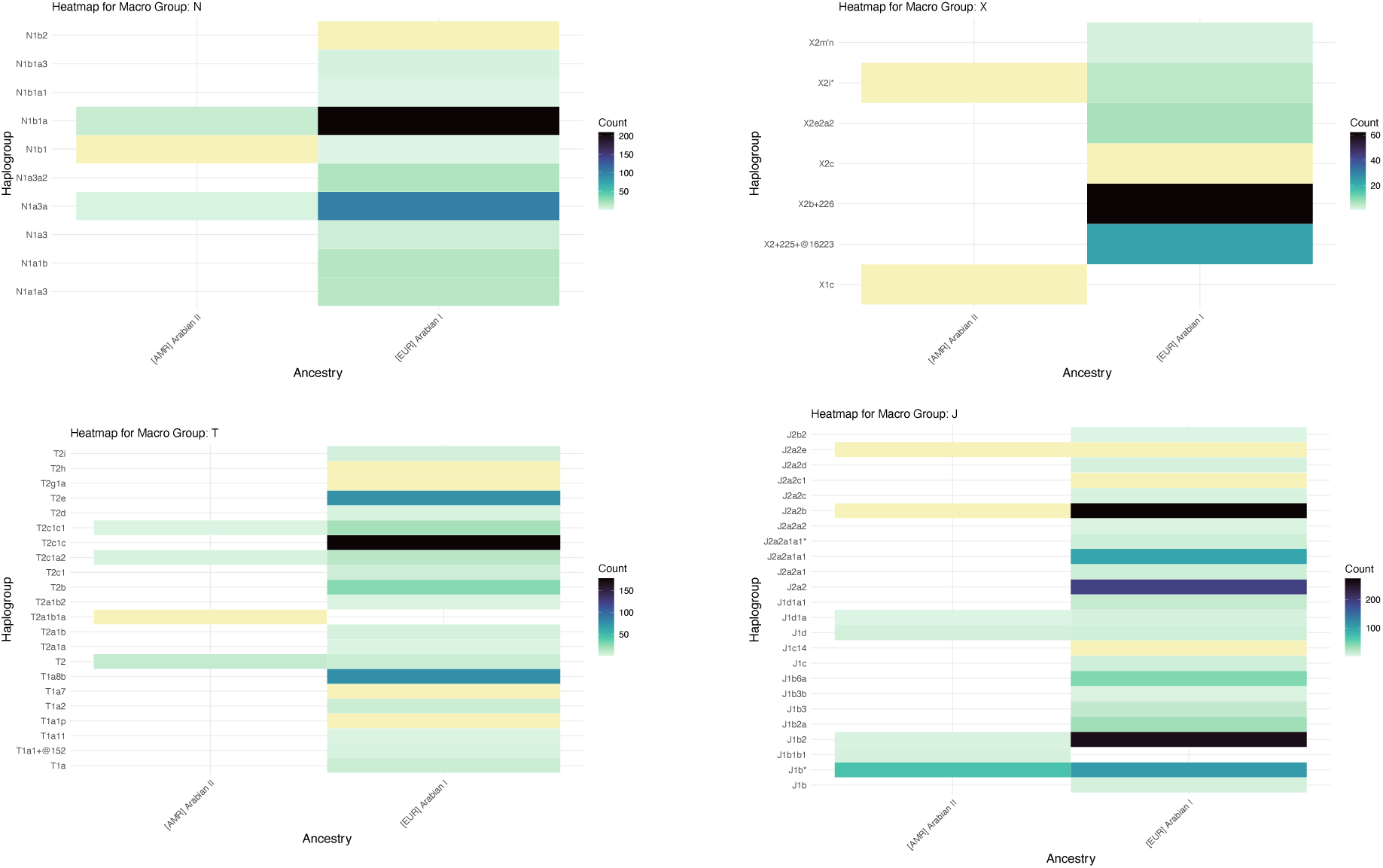
Mitochondrial haplogroups of macrohaplogroups X, N, T and J detected in EGP individuals with more than 90% of the respective admixture component. The color code denotes how many of them carry a given haplogroup.

**Supplementary Figure 14.**
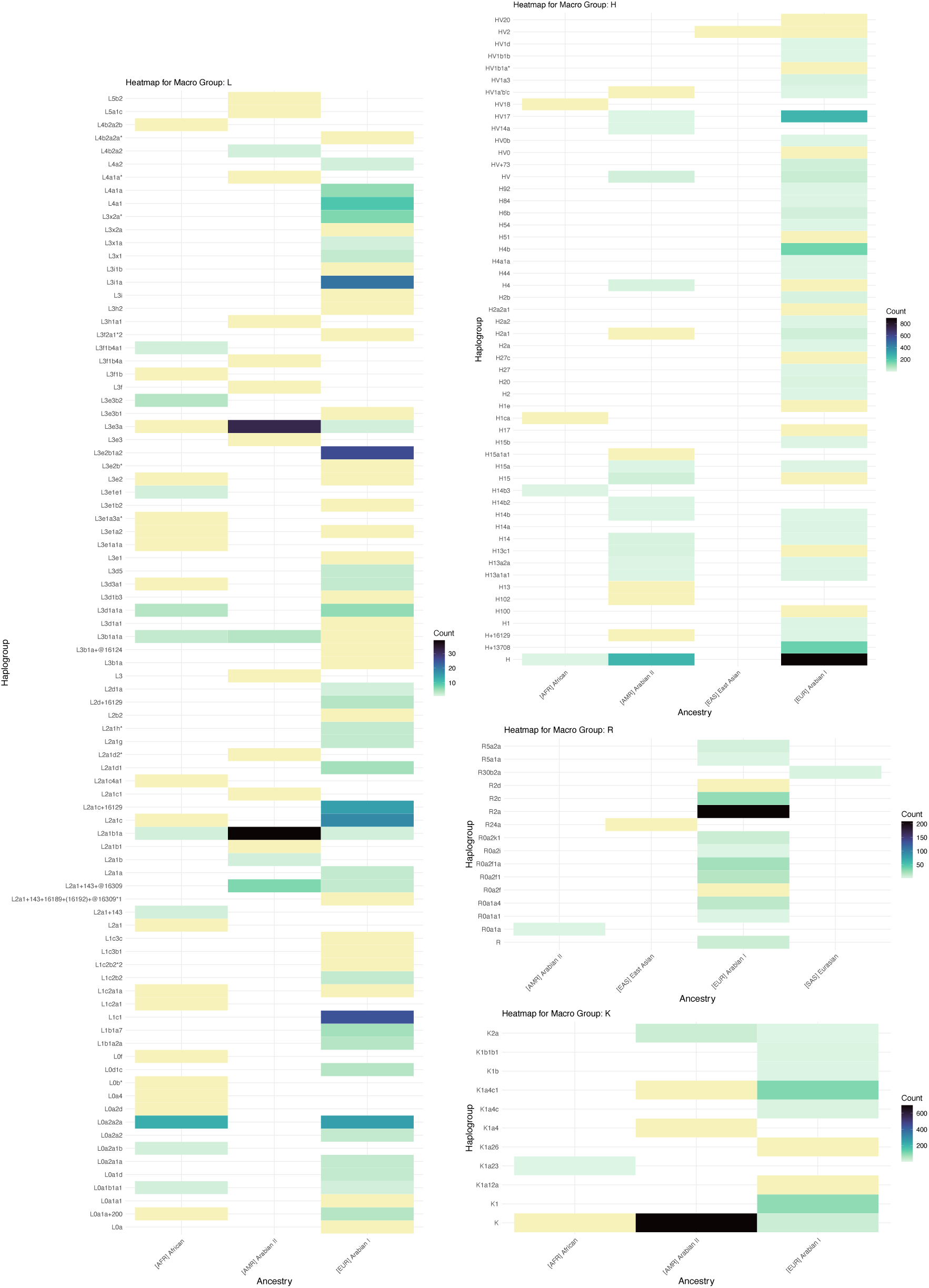
Mitochondrial haplogroups of macrohaplogroups L, H, R and K detected in EGP individuals with more than 90% of the respective admixture component. The color code denotes how many of them carry a given haplogroup.

**Supplementary Figure 15.**
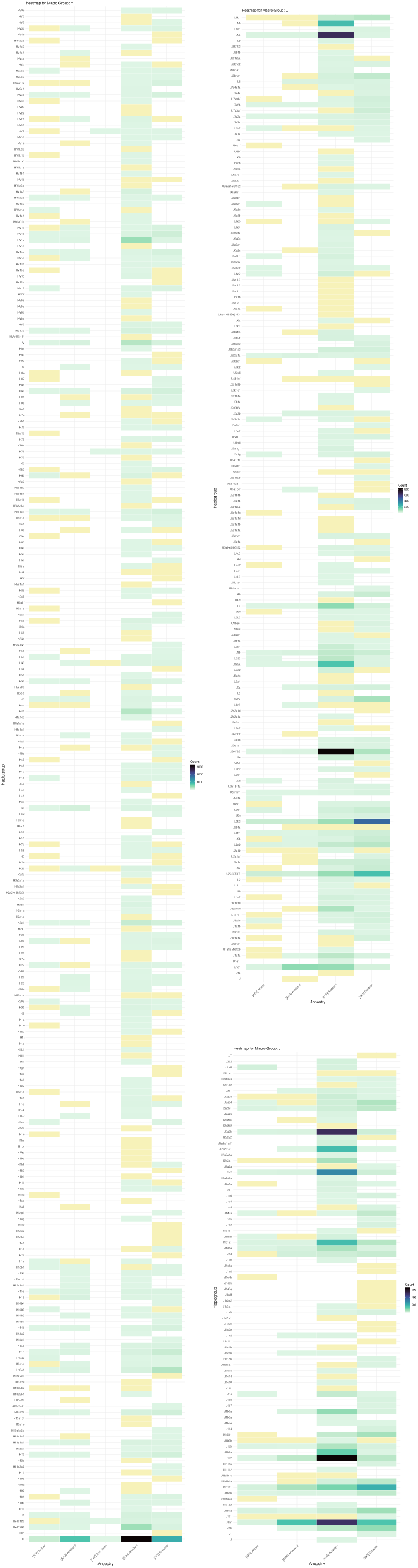
Mitochondrial haplogroups of macrohaplogroups H, J and U detected in EGP, stratified by main admixture component. The color code denotes the number of individuals with a given admixture component who carry a given haplogroup.

**Supplementary Figure 16.**
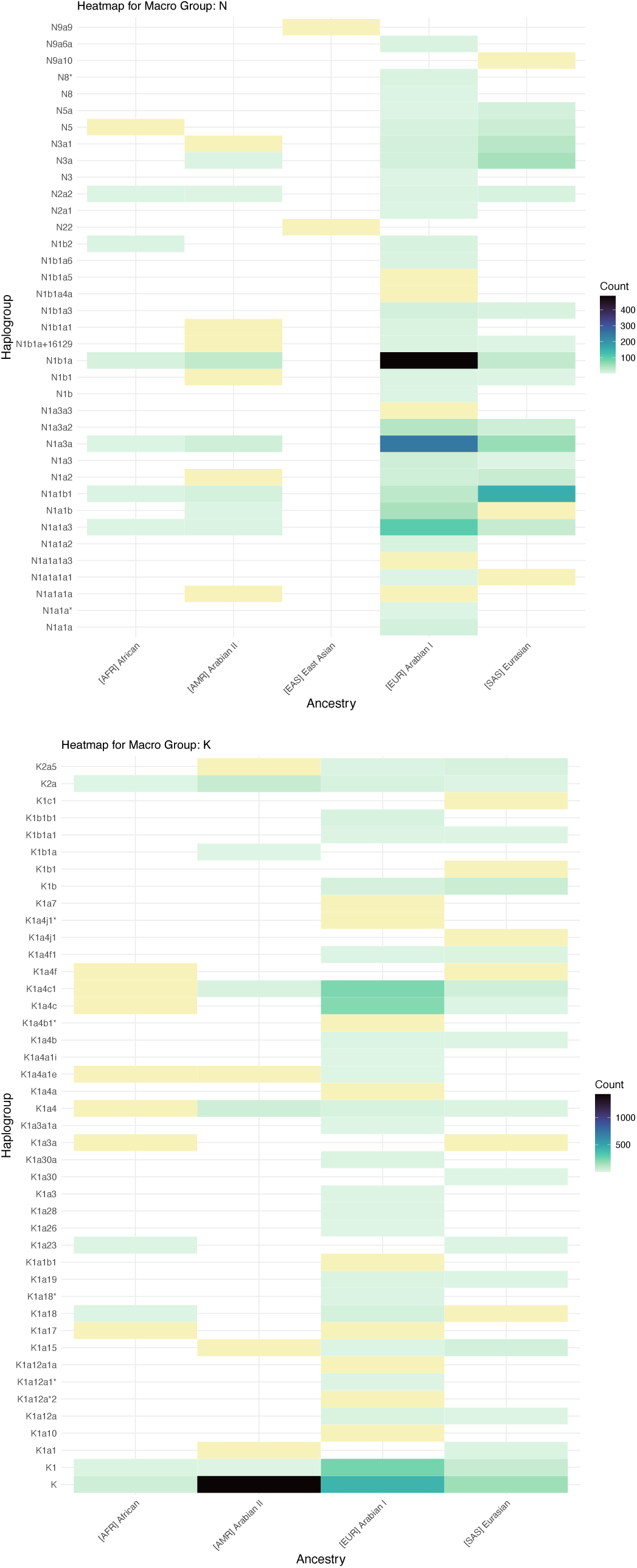
Mitochondrial haplogroups of macrohaplogroups K and N detected in EGP, stratified by main admixture component. The color code denotes the number of individuals with a given main admixture component who carry a given haplogroup.

**Supplementary Figure 17.**
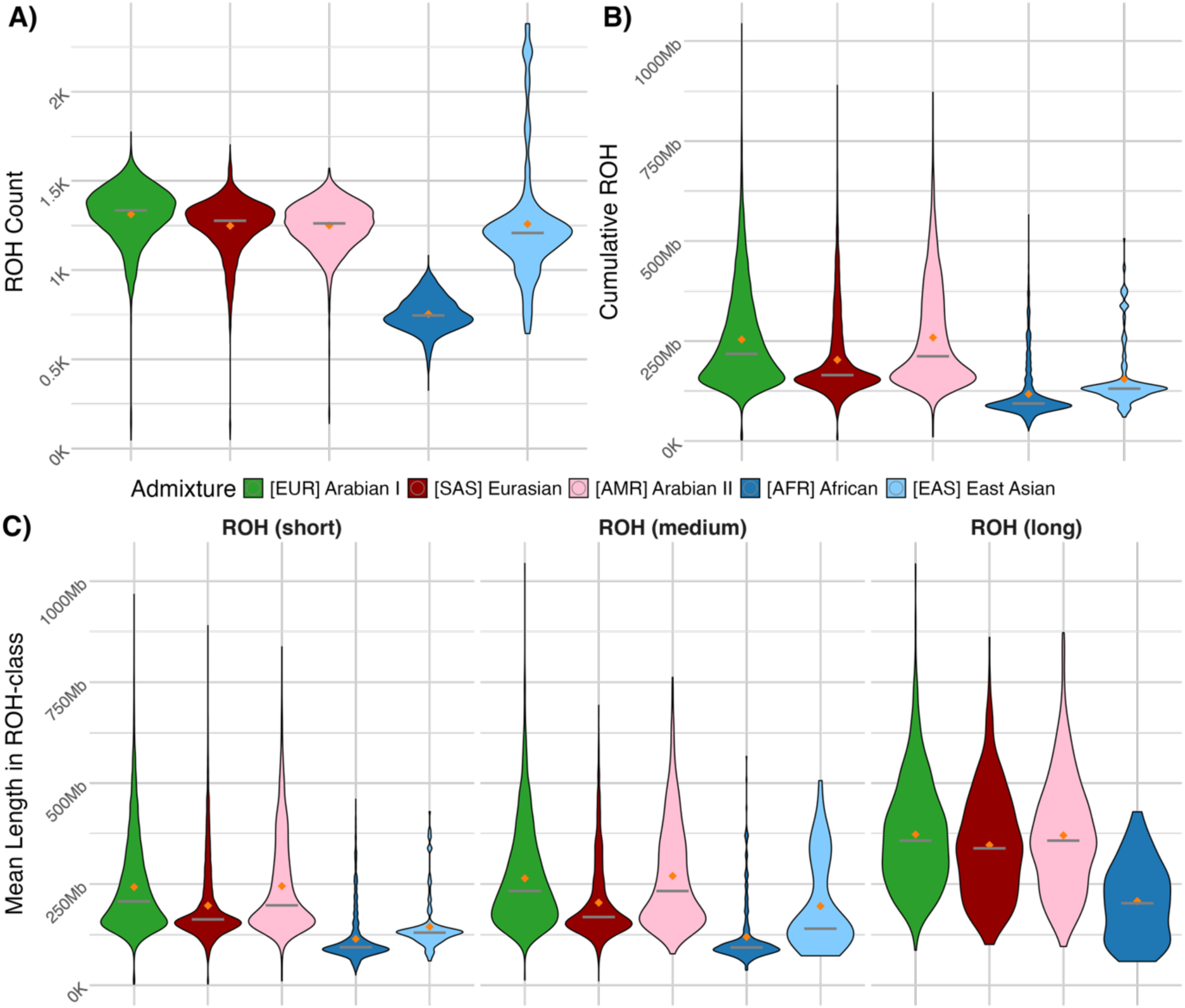
Panels A) and B) depict the count and total length of ROHs in each sample stratified by the five main ancestry components, respectively. Significant variations are observed across ancestral components, notably in the mean number of ROHs. The [AFR] African component exhibits the lowest mean number of ROHs. In contrast, [EUR] Arabian I, positioned proximal to European origins, displays the highest mean number of ROHs. Panel C) shows the Gaussian Mixture Model Clustering of Runs of Homozygosity (ROH) Metrics Lengths into Three Length Categories. This figure illustrates the clustering of Runs of Homozygosity (ROH) into three distinct length categories using a Gaussian Mixture Model. The resulting categories are defined as short (ROH length < 14.73 Mb), medium (ROH length > 14.73 Mb and < 63.03 Mb), and long (ROH length > 63.03 Mb).

**Supplementary Figure 18.**
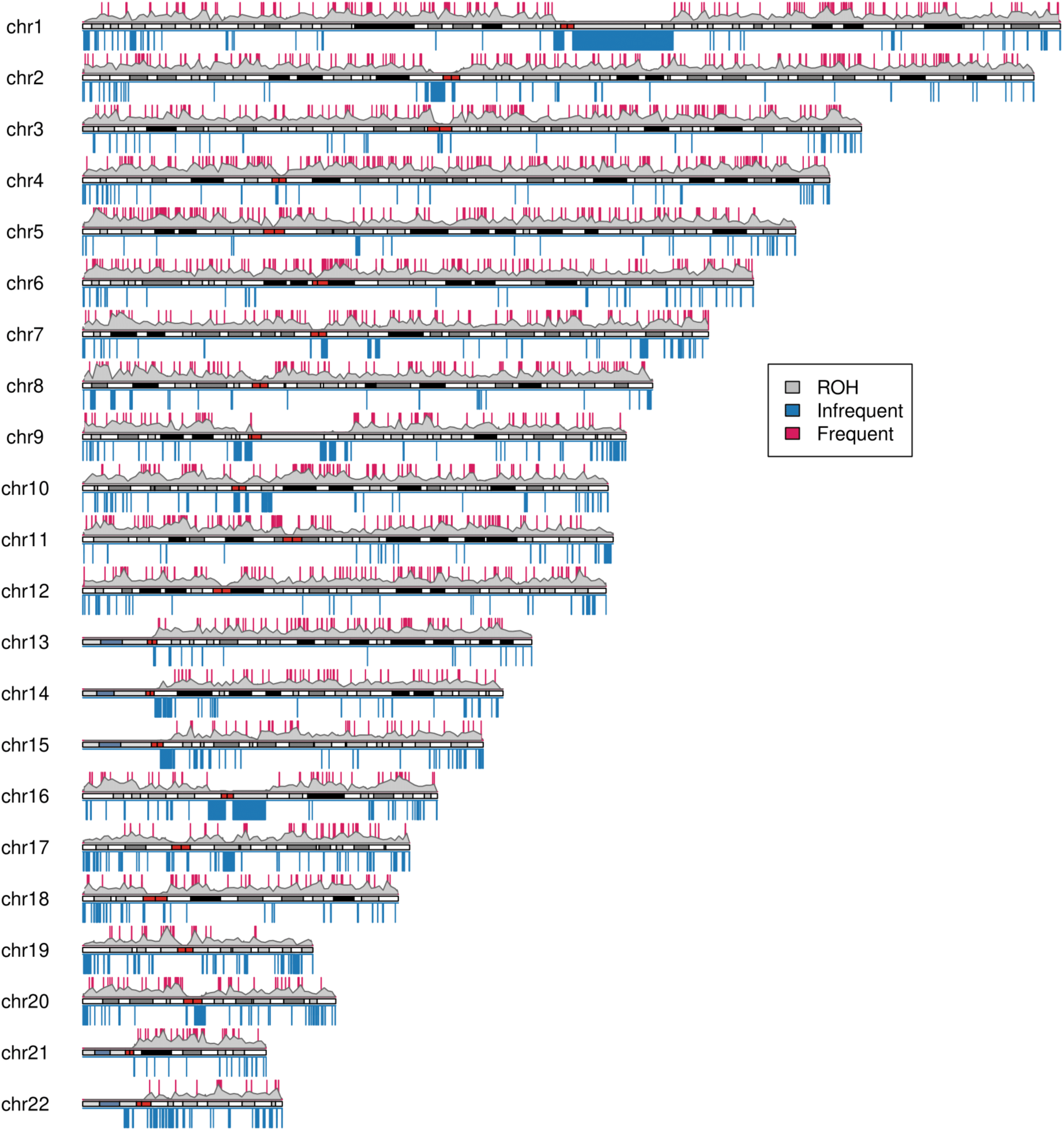
Ideogram Illustrating the Distribution of Runs of Homozygosity (ROH) on top of each autosome. This plot provides an overview of the cohort-wide density distribution of Runs of Homozygosity (ROH) across each autosome. A window size of 10 kb was used to calculate the median number of samples in adjacent regions. The ROH regions presented in this plot have been filtered based on a minimum/maximum prevalence criterion, with 5 percent quantile (1500 samples) and 95 percent quantile (8029 samples) cutoffs applied. Centromere regions were removed from the plot. Regions meeting the five percent prevalence are denoted in blue below each chromosome representation, indicating relatively infrequent occurrences. Conversely, regions surpassing the 95 percent prevalence are highlighted in red, signifying high-frequency ROH regions. The enrichment analyses revealed no significant biological function enrichment in either the frequent or infrequent regions.

**Supplementary Figure 19.**
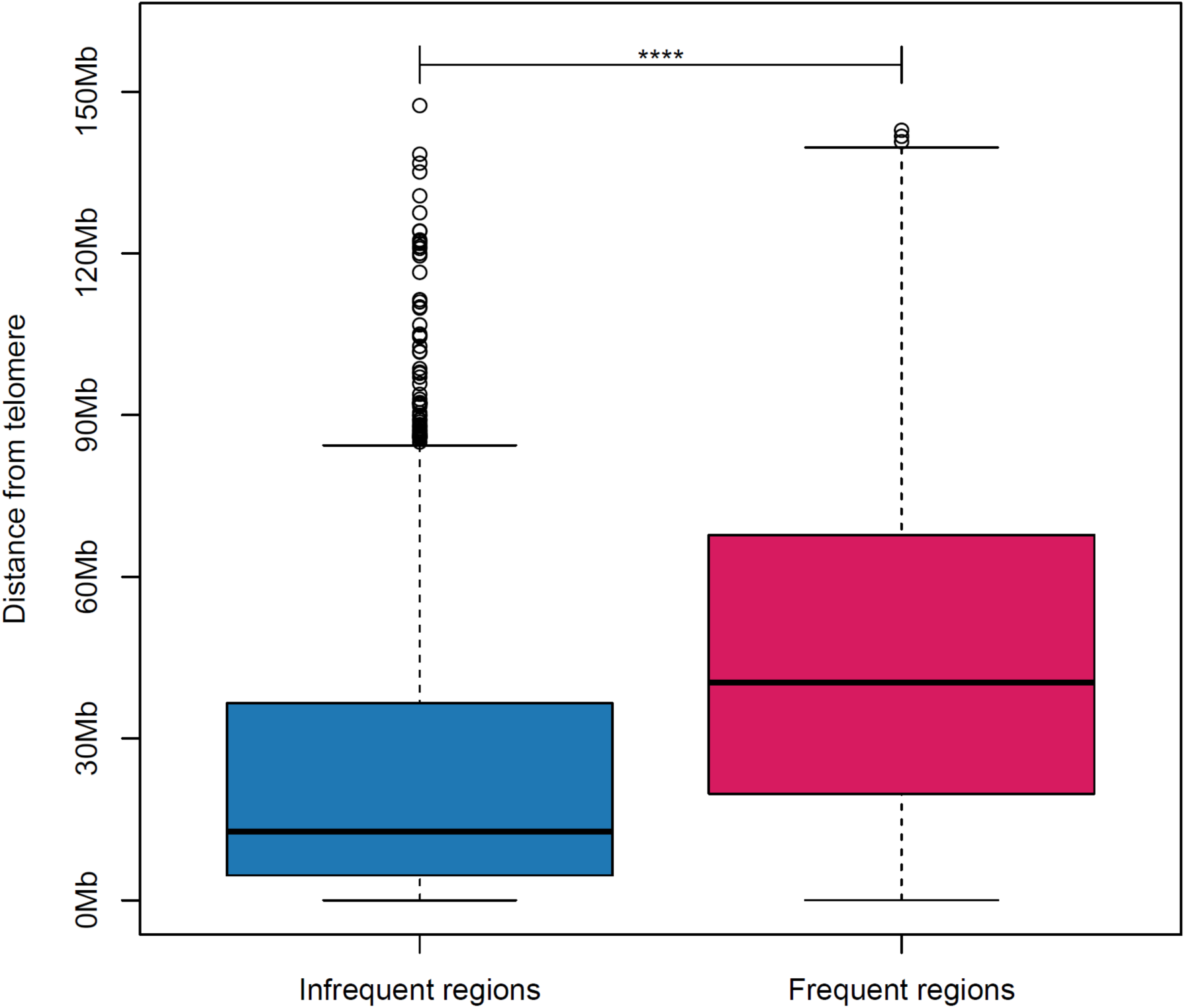
Boxplot shows the difference in proximity to the telomere between infrequent and frequent ROHs. Genomic regions were divided into 10Kb windows, and the number of samples with ROH in each window was registered. Frequent ROHs are defined as the top 5% in frequency, whereas the bottom 5% are categorized as infrequent ROHs. This distance measurement is based on the span from the ROH to the telomere within the same chromosome arm, and adjacent windows were treated as a single region. Infrequent ROHs are significantly closer to the telomere than frequent ROHs. The statistical comparison was performed using a t-test. Four asterisks (****) represent p < 0.0001.

**Supplementary Figure 20.**
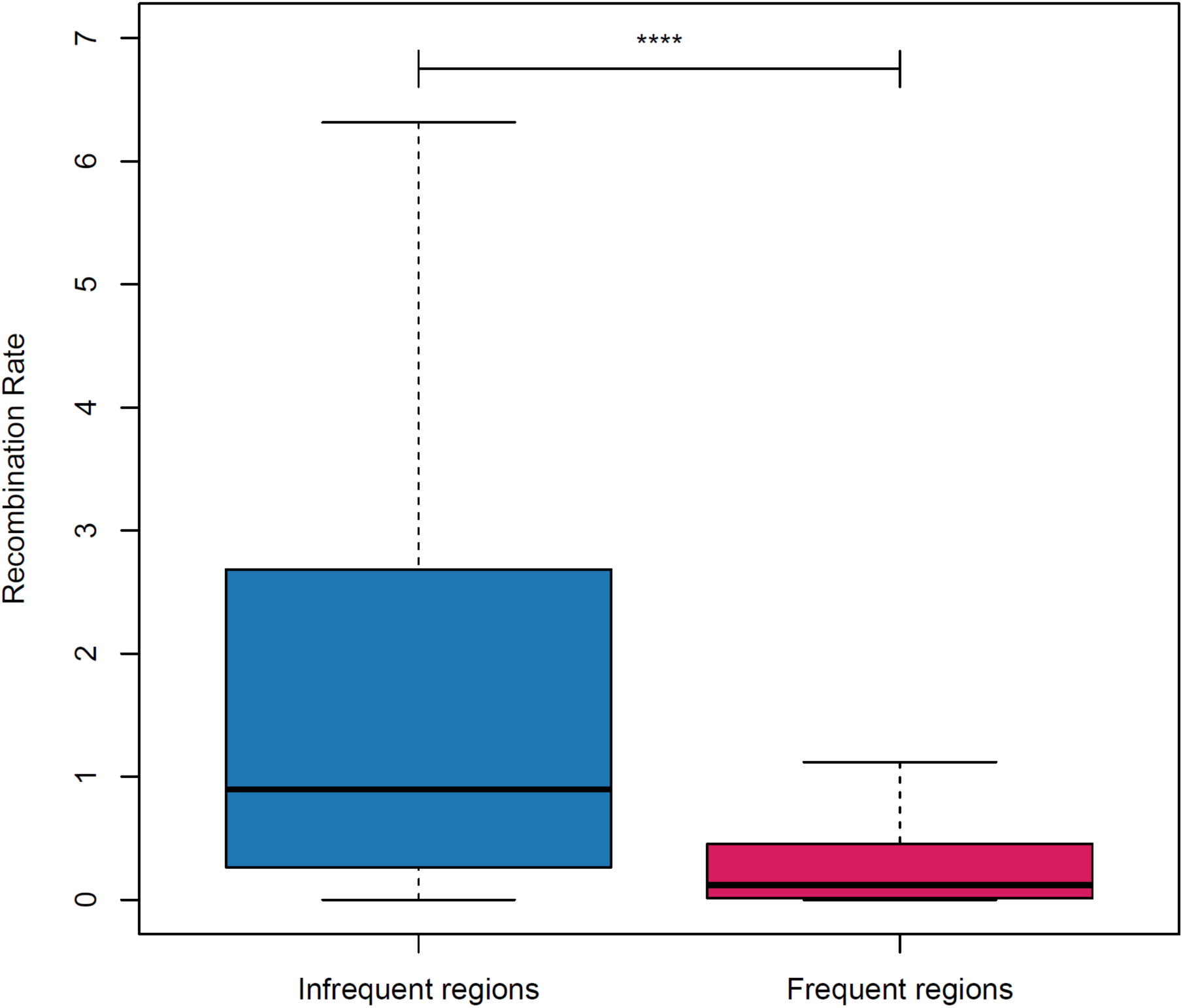
Boxplot shows the difference in the recombination rate of infrequent and frequent ROHs. Genomic regions were divided into 10 kb windows, and the number of samples with ROH in each window was registered. Frequent ROHs are defined as the top 5% in frequency, whereas the bottom 5% are categorized as infrequent ROHs. The average recombination rate from the 1,000 Genomes Project was assigned to each window labeled as infrequent and frequent. Infrequent ROHs have a significantly higher recombination rate than frequent ROHs. The statistical comparison was performed using a t-test. Four asterisks (****) represent p < 0.0001.

**Supplementary Figure 21.**
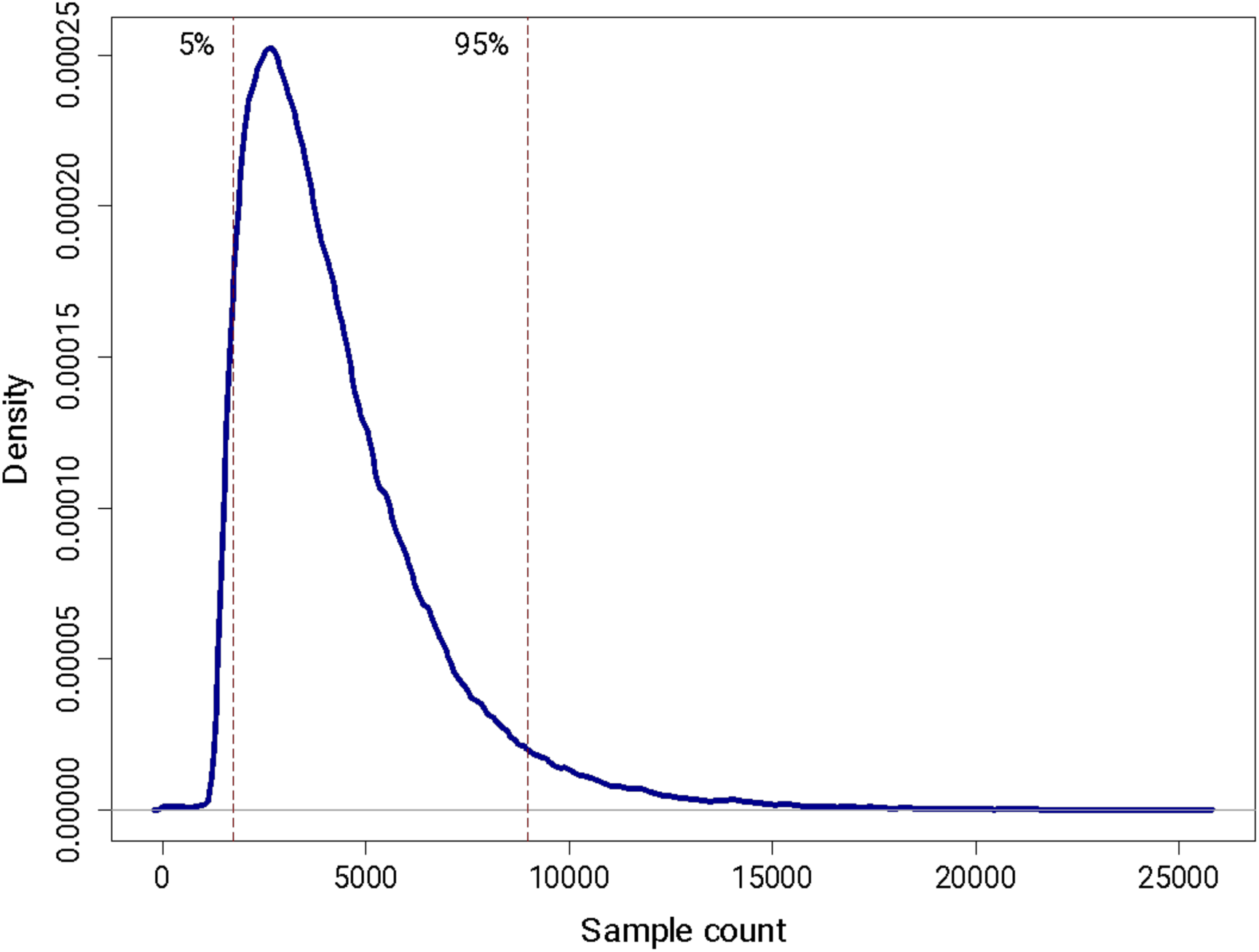
Distribution of Runs of Homozygosity (ROH) Shared by Multiple Samples. The 5th and 95th percentiles indicate the cutoffs for the regions’ lowest and highest 5%, respectively. The vertical axis shows the frequency of shared ROHs, while the horizontal axis represents the number of samples. The number of samples sharing a given region (10 kb window size) was calculated using the kpPlotDensity function from the karyoploteR package in R. The 95% quantile (8,029 samples) and the 5% quantile (1,500 samples) were used to determine the cutoffs for the highest- and lowest-frequency regions, ensuring an even distribution of frequent and infrequent ROHs. Only autosomes are included in this analysis. Centromere regions were excluded from the analysis.

**Supplementary Figure 22.**
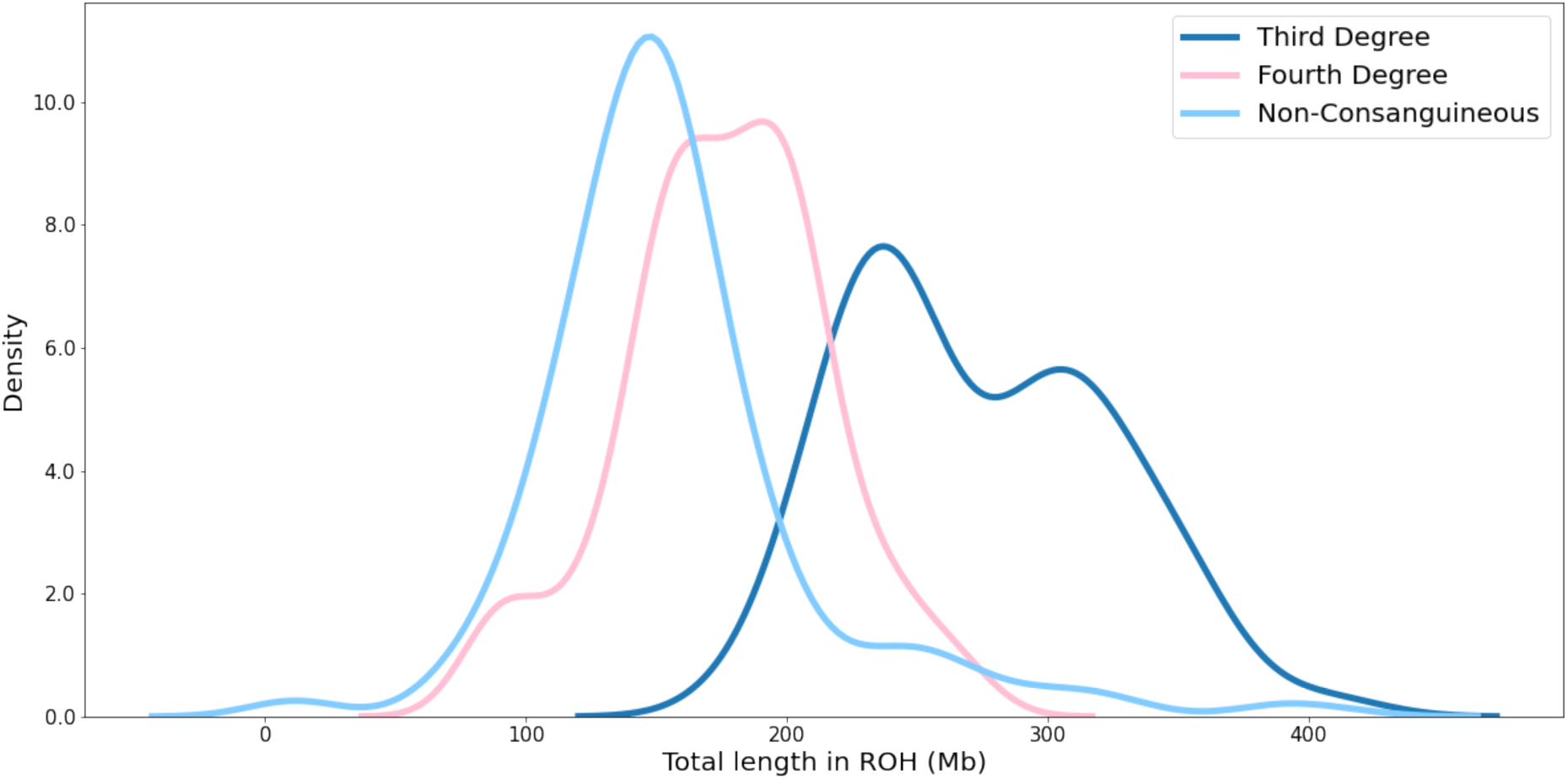
Density Plot Illustrating the Total Length of Runs of Homozygosity (ROH) Metrics in Offspring. This figure showcases a density plot depicting the distribution of the total length of Runs of Homozygosity (ROH) in offspring, categorized based on the inferred degree of non-/consanguinity of parents. The density plot presents a nuanced view of how the total length of ROH varies across offspring groups with differing degrees of parental relatedness. Parental non-/consanguinity is inferred based on genetic data, allowing classification into various degrees of relatedness. The density plot visually captures these distinctions, offering insights into the genetic landscape associated with different levels of parental kinship. This visualization contributes to a comprehensive understanding of the distribution of ROH lengths among offspring, providing valuable insights into the genetic implications of parental non-/consanguinity. In this context, Third Degrees are those with a pi-hat more than 0.125. Fourth Degree is that with a pi-hat between 0.0625 and 0.125. Non-consanguineous are those with a pi-hat value below 0.0625.

**Supplementary Figure 23.**
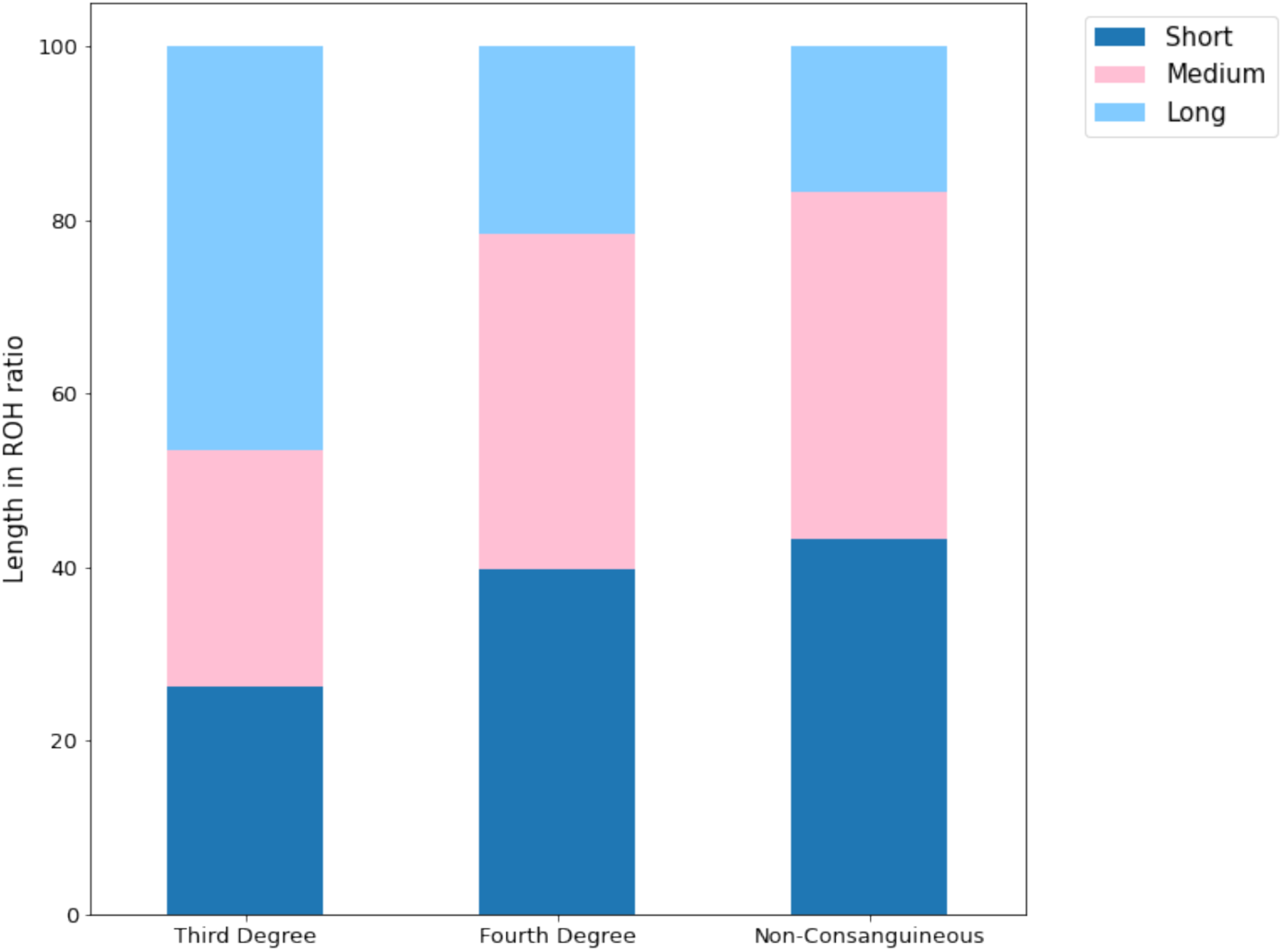
Runs of Homozygosity (ROH) Length Gaussian Clustering for the Cohort. This figure depicts the Gaussian Mixture Model clustering of Runs of Homozygosity (ROH) total lengths stratified by hereditary groups. The stacked barplot shows the ratio of region length categories averaged over all samples in the respective group. This figure analysis encompasses data from 511 (3rd-degree: 60, 4th-degree: 187, non-consanguineous: 264) individuals, offering a robust and representative examination of ROH patterns within the studied cohort.

**Supplementary Figure 24.**
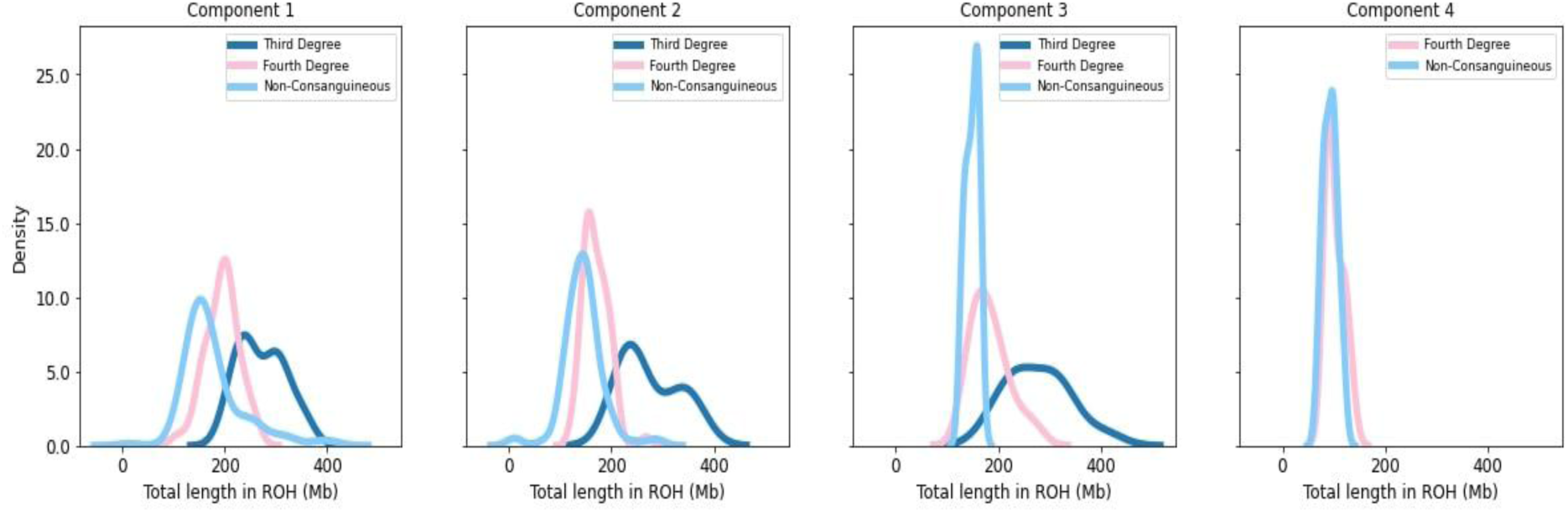
Density Plot Illustrating the Total Length of Runs of Homozygosity (ROH) Metrics in Offspring, stratified by admixture components. This figure presents a density plot showcasing the distribution of the total length of Runs of Homozygosity (ROH) in offspring. The data is stratified based on the inferred degree of non-/consanguinity of parents, further split to account for detected admixture components. It is worth noting that, unfortunately, complete families were not available for the fifth main ancestry component. The density plot provides a comprehensive visual representation, capturing the variability in the total length of ROH among offspring groups. Stratification based on parental non-/consanguinity and further division by detected admixture components enriches our understanding of the genetic landscape. Complete families for the fifth main ancestry component were not available. This limitation underscores the necessity of considering data availability when interpreting results related to this specific component.

**Supplementary Figure 25.**
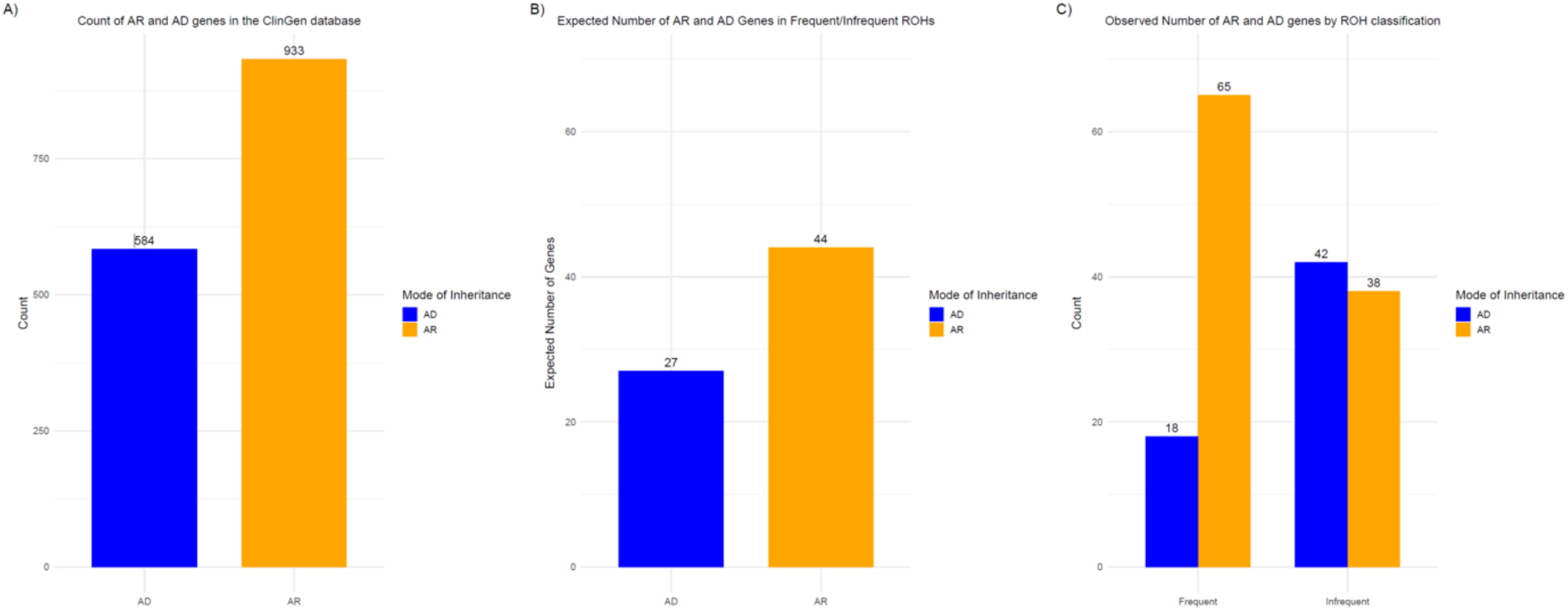
Distribution of Autosomal Recessive (AR) and Autosomal Dominant (AD). **A)** The total count of AR and AD genes in the ClinGen database, where 933 genes are classified as AR, and 584 as AD. These genes have definitive disease-gene associations on autosomes (chr1-22), based on ClinGen annotations as of December 5, 2024 (Supplementary Table 16). **B)** Expected number of AD and AR genes in frequent and infrequent ROHs. Considering that the total lengths of frequent and infrequent ROHs are nearly identical (∼136 MB), representing approximately 4.7% of the autosomes (136 MB / 2904 MB, hg38), the expected number of AR and AD genes in these ROH regions can be calculated. The expected number of AR genes is approximately 44, calculated as 4.7% * 933 AR genes, and the expected number of AD genes is around 27, calculated as 4.7% * 584 AD genes. **C)** Number of AR and AD genes overlap the frequent and infrequent ROH regions identified in this study (Supplementary Table 15). The observed number of genes in these ROH regions deviates from the expected values, showing a higher enrichment of AR genes in the frequent ROHs and AD genes in the infrequent ROHs, with a lower enrichment of AD genes in the frequent ROHs. This suggests distinct patterns of genetic variation with important implications for population-level genetic studies.

**Supplementary Figure 26.**
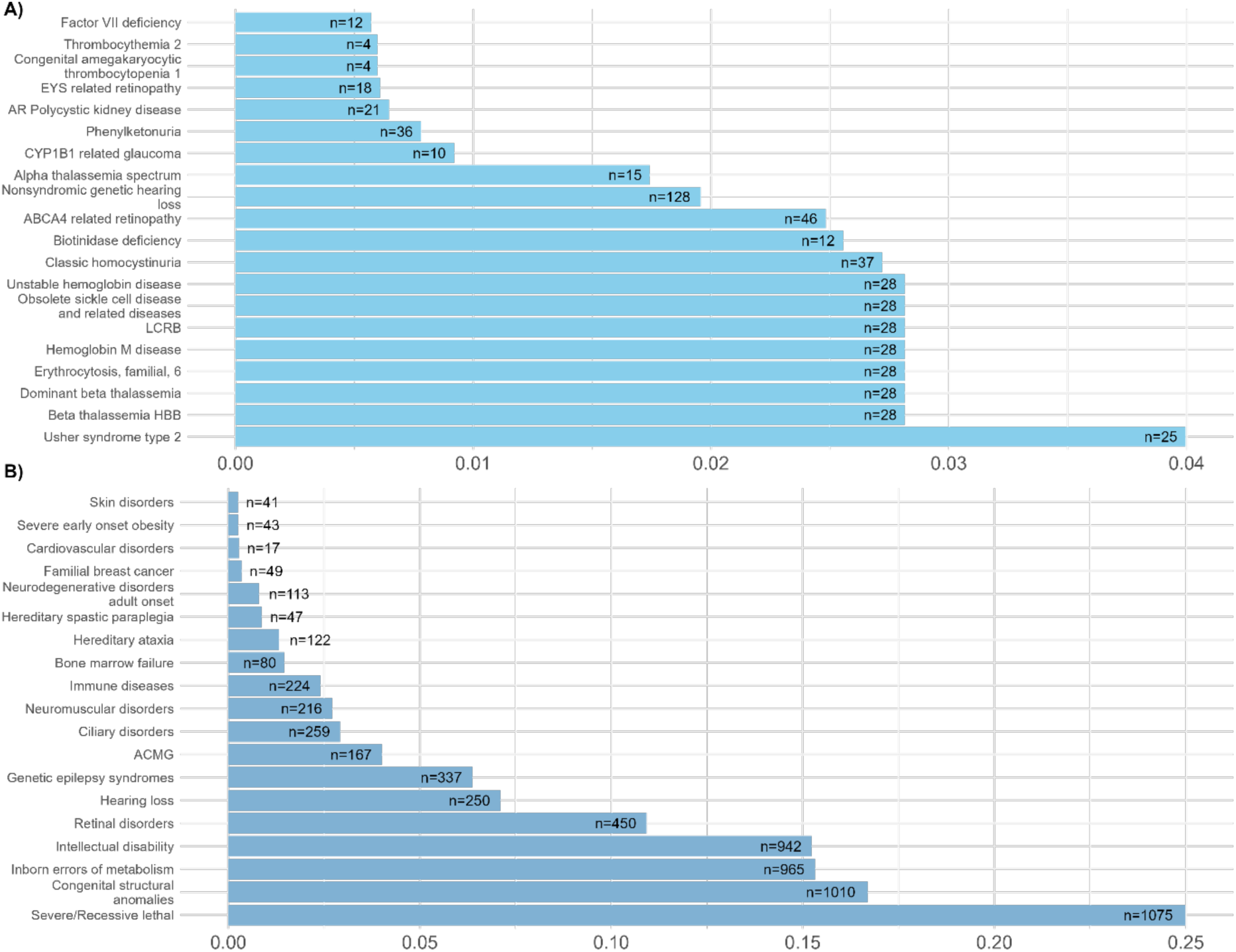
Cumulative Allele Frequency of Pathogenic/Likely Pathogenic Variants in Mendelian Inherited Diseases and Disease Categories. This figure displays the cumulative allele frequency (AF) of pathogenic (P) and likely pathogenic (LP) variants in Mendelian inherited diseases and disease categories in the UAE cohort. Panel **A** shows the top 20 diseases with the highest cumulative AF of P/LP variants, with the x-axis representing the cumulative AF of P/LP variants and the number of observed P/LP variants (n) indicated for each disease. The diseases are linked to the genes harboring the variants based on established gene-disease relationships rather than direct variant-disease causality. Panel **B** shows the cumulative AF of P/LP variants in different disease categories (n=19), with the x-axis representing the cumulative AF of P/LP variants and the number of observed P/LP variants (n) indicated for each category. The full list of diseases and disease categories and their corresponding data are presented in Supplementary Tables 17 and 18, respectively.

**Supplementary Figure 27.**
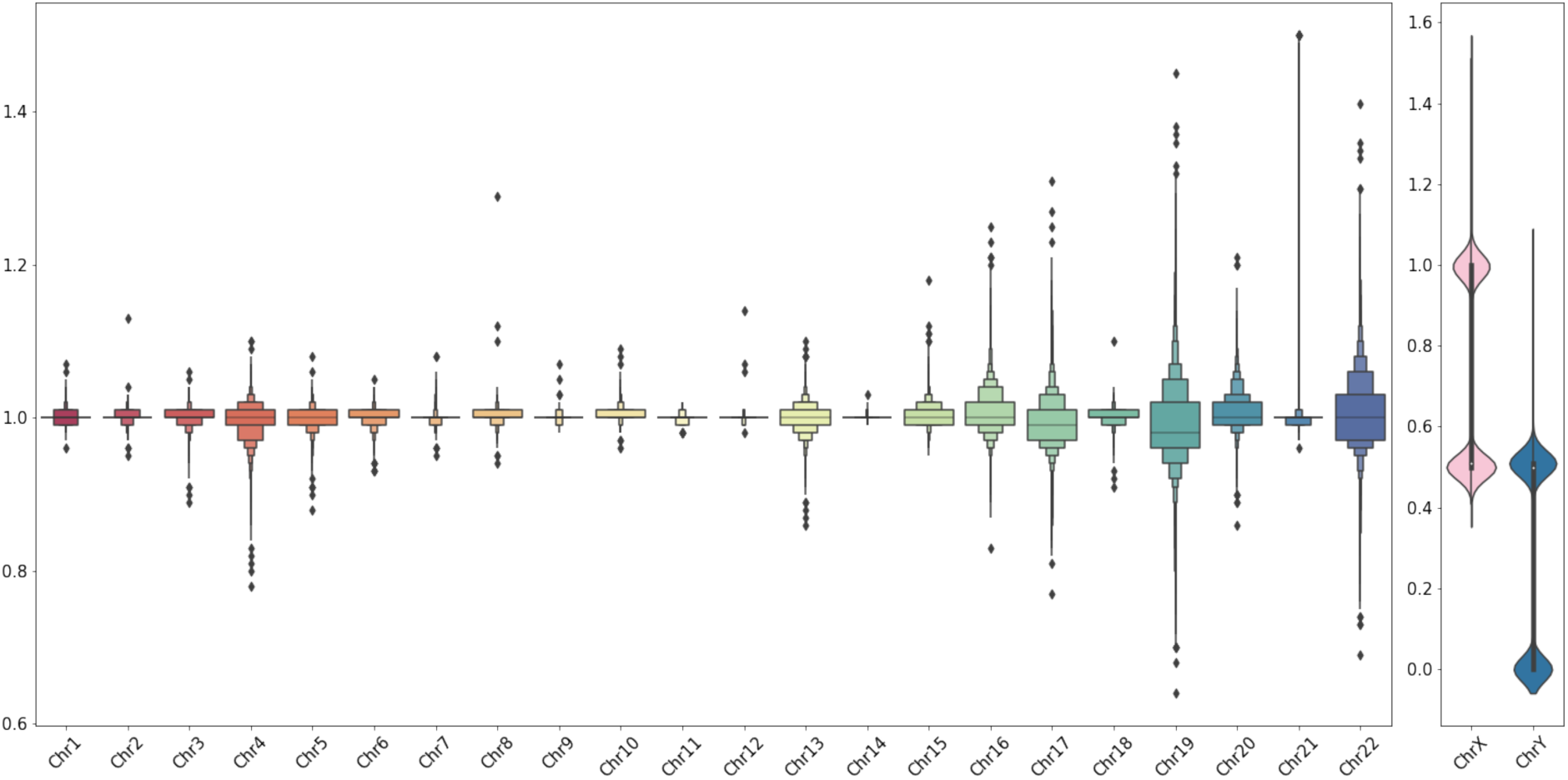
Boxplot illustrating ploidy estimation coverage across autosomes and sex chromosomes. The anticipated ploidy value for autosomes is one, denoting the presence of both chromosome copies in a sample. In males, the expected ploidy value for the Y chromosome is 0.5, while in females, it is zero. Minor deviations from these values are typically anticipated. However, a ploidy value below 0.75 may suggest the absence of one chromosome copy in a sample.

**Supplementary Figure 28.**
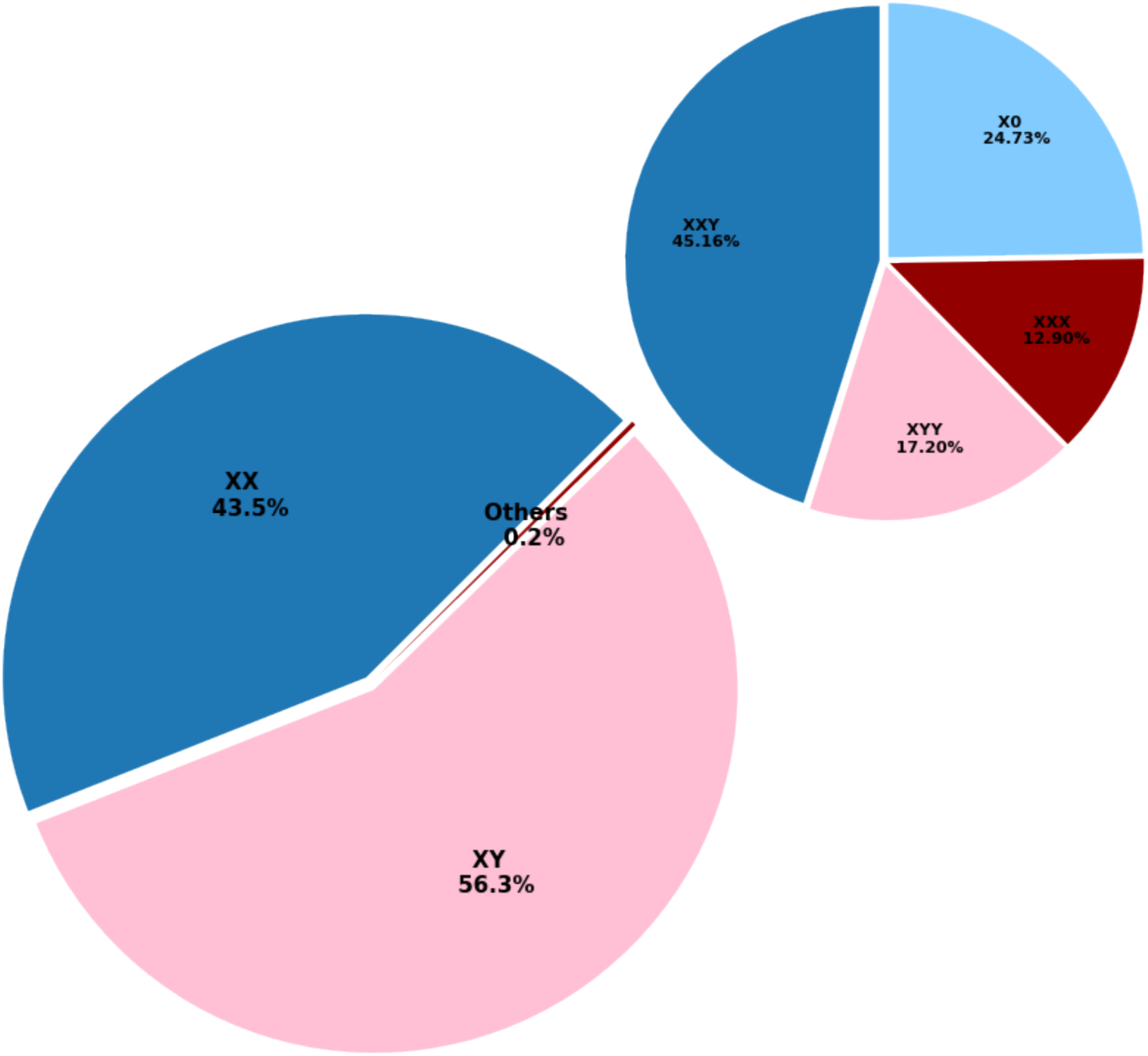
Distribution of Sex Chromosome Abnormalities in the Cohort. This figure illustrates the prevalence of sex chromosome abnormalities within the cohort. Among the participants, 93 samples exhibit anomalies in sex chromosomes. The identified abnormalities are categorized into distinct types, including XXX, XXY, XYY, and X0. One case in which DRAGEN could not determine the sex chromosome ploidy reliably was most compatible with an XXY configuration and is counted as such here. The observed ratio of 0.2% within the population aligns closely with the reported worldwide average rate.

**Supplementary Figure 29.**
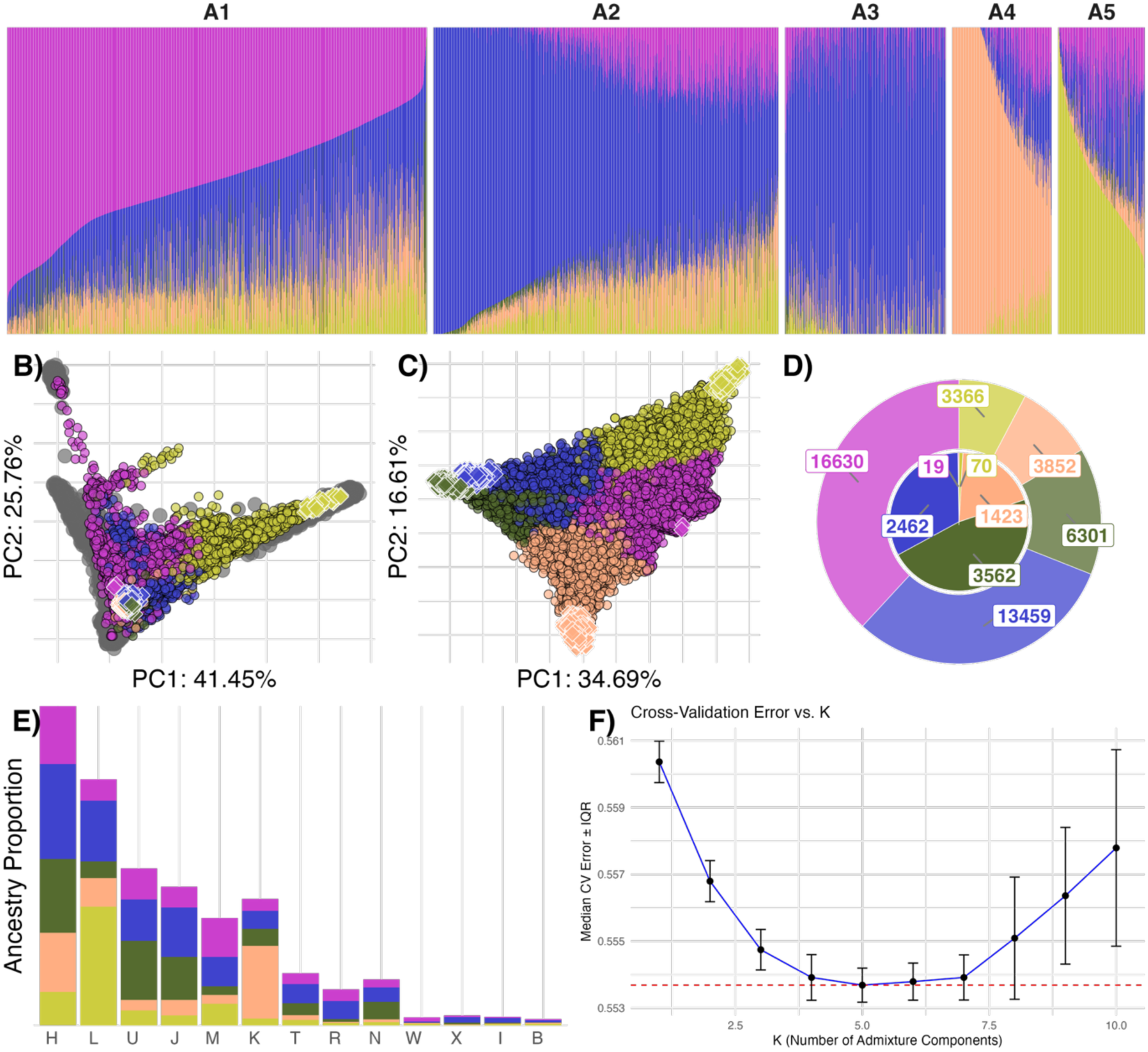
Genetic stratification of the study cohort based on unsupervised admixture and principal component analyses. Unsupervised admixture analysis was performed using K = 5, and each sample was assigned a primary ancestry component, defined as the component with the highest contribution. A) Bar plots showing ancestry component proportions for all individuals grouped by their primary ancestry component. Within each group, samples are ordered by decreasing the proportion of the primary component. B) Principal Component Analysis (PCA) of the cohort projected onto axes defined by the 1000 Genomes (G1K) reference dataset. Samples are colored by primary ancestry component (black outline), and those with >90% contribution from a single component are highlighted as diamonds (white outline). C) PCA projection of G1K reference samples (black outline) onto axes defined by the ERGP cohort. ERGP samples with >90% primary ancestry component are again highlighted as diamonds (white outline). D) Pie chart showing the distribution of primary ancestry components across the full cohort (outer ring) and for samples with >90% primary component contribution (inner ring). E) Distribution of mitochondrial haplotypes stratified by primary ancestry component, sorted by set size. F) Elbow plot showing median cross-validation (CV) error across 100 replicates of 1,000 randomly sampled individuals. The minimum median error is observed at K = 5, supporting the selected model complexity.

